# Causes of stillbirth in sub-Saharan Africa and South Asia: Findings from Child Health and Mortality Prevention Surveillance, 2016-2023

**DOI:** 10.1101/2025.09.09.25335469

**Authors:** Afruna Rahman, Kyu Han Lee, Shams El Arifeen, Mohammad Zahid Hossain, Atique Iqbal Chowdhury, Muntasir Alam, Afsana Afrin, Ikechukwu Udo Ogbuanu, Solomon Samura, Erick Kaluma, Julius Ojulong, Samba Sow, Adama Mamby Keita, Milagritos D. Tapia, Kiranpreet Kaur Chawla, Karen Kotloff, Shabir A. Madhi, Sana Mahtab, Yasmin Adam, Amy Wise, Ziyaad Dangor, Christopher Mugah, Elizabeth Oele, Richard Omore, Aggrey K. Igunza, Dickens Onyango, Inacio Mandomando, Quique Bassat, Sara Ajanovic, Rosauro Varo, Elisio Xerinda, Nega Assefa, J. Anthony G. Scott, Lola Madrid, Melisachew M Yeshi, Fikremelekot Temesgen, Lucy Liu, Portia Mutevedzi, Dianna M. Blau, Victor Akelo, Robert F. Breiman, Cynthia G. Whitney, Emily S. Gurley, the CHAMPS Consortium

## Abstract

**Background:** Globally, an estimated 1.9 million stillbirths occur annually, yet significant knowledge gaps exist regarding the causes of stillbirths, particularly in high-burden regions. We investigated fetal and maternal conditions causing stillbirths in seven countries throughout sub-Saharan Africa and South Asia and described missed opportunities for prevention.

**Methods:** Child Health and Mortality Prevention Surveillance (CHAMPS) identified stillbirths at sites in Bangladesh, Ethiopia, Kenya, Mali, Mozambique, Sierra Leone, and South Africa. We asked families for consent to conduct minimally invasive tissue sampling (MITS) from December 2016 to December 2023. An expert panel reviewed test results, clinical information, and verbal autopsy data and assigned fetal and maternal conditions leading to stillbirths and identified missed opportunities for prevention.

**Findings:** A cause of death (fetal or maternal condition) was determined for 94% (2342/2492) of stillbirths; the most frequent condition in the fetus was intrauterine hypoxia (75%, 1864/2492) across all sites, resulting from either maternal conditions (49%, 922/1864) or placental causes (33%, 610/1864). Congenital infections were determined to be the cause of 9% (228/2492) of stillbirths, accounting for the highest proportion in South Africa (28%, 87/306). Group B Streptococcus, *Escherichia coli*, and *Enterococcus faecalis* were the most common causative pathogens. Congenital birth defects caused 9% (227/2492) overall and were most common in Ethiopia (24%, 134/568). Primary maternal conditions were identified in 72%, most often placental complications (18%, 446/2492) and maternal hypertension (17%, 414/2492). Placental complications were more common in Mali (43%, 92/212) while maternal medical and surgical conditions were most frequently observed in South Africa (40%, 121/306) and Bangladesh (39%, 158/405). Most (72%, 1808/2492) causes of stillbirth were considered preventable, with heterogeneity observed across sites on the recommended prevention strategies.

**Interpretation:** Complications of pregnancy or delivery were responsible for a large majority of stillbirths. Among the fetal conditions identified, infections and congenital defects were the most common. This study identified widespread gaps in antenatal care and obstetric services as the main drivers of stillbirths. However, there was considerable geographic heterogeneity in underlying causes and recommended prevention measures, suggesting that strategies to reduce stillbirths should be informed by local data to be optimally successful.

**Funding:** The Gates Foundation

## Introduction

Five stillbirths occur each minute, leading to an estimated global total of 1.9 million in 2021; 98% occur in low- and middle-income countries (LMICs).^1^ In sub-Saharan Africa and South Asia, third-trimester stillbirth rates are nearly tenfold higher than in high-income countries.^2^ Despite a decline in global stillbirth rates over the past two decades, progress lags behind infant and child survival improvements, falling short of global targets.^1^ In 2021, in sub-Saharan Africa and South Asia, the estimated stillbirth rate was 21.1 and 17 per 1000 total births, respectively.^1^ The World Health Organization (WHO) Every Newborn Action Plan aimed to lower global stillbirth rates to 12 or fewer per 1,000 live births by 2030.^3^ To reach this goal, interventions must prioritize common causes of stillbirths, particularly in high-burden areas.^1,3^

The dearth of quality data from LMICs on the etiology of stillbirths poses a significant challenge not only in constructing accurate statistics around the specific causes of fetal deaths but also in designing programs to prevent stillbirths. Our present understanding of stillbirths primarily stems from vital registration data and verbal autopsies.^4^ However, both tools have substantial limitations, particularly when inferring etiologic causes. Many LMICs do not include stillbirths in vital registration systems, and verbal autopsies for stillbirths consist of only a few questions related to family reports of the events surrounding the stillbirth.^4^ Neither approach involves a review of maternal medical records or a systematic biological investigation of the fetus and placenta, processes that are essential for determining causes of stillbirth.^5^ A systematic review from 2018 showed that studies inconsistently applied classification systems for causes of stillbirth and suffered from poor-quality data, resulting in findings that were insufficient to guide prevention strategies.^6^ In many countries, policies for identifying and documenting stillbirths are less developed compared to those for neonatal and under-five deaths. This gap likely contributes to the slow progress in reducing stillbirths, as inadequate data collection hinders targeted prevention and intervention efforts.

The Child Health and Mortality Prevention Surveillance (CHAMPS) Network aimed to track stillbirth and under-five mortality causes in South Asia and sub-Saharan Africa—regions with the highest stillbirth rates—using robust, standardized longitudinal data.^9^ Early findings from the first two years of CHAMPS showed that intrauterine hypoxia was the main fetal cause of death, contributing to 72% of 180 stillbirths investigated using minimally invasive tissue sampling (MITS).^10^ Similar findings were reported in the PURPOSe study, which examined 611 stillbirths in India and Pakistan; however, both studies had major limitations for informing global prevention strategies, either due to small sample sizes or limited geographic scope. In this paper we analyzed the placental, maternal, and fetal conditions leading to 2,492 stillbirths across seven CHAMPS countries in Sub-Saharan Africa and South Asia from 2016-2023, including identifying prevention opportunities missed.

## Methods

### Sites and mortality surveillance

A comprehensive description of CHAMPS sites and standardized mortality surveillance methods are available elsewhere.^11^ Briefly, the CHAMPS Network consists of sites in seven countries in sub-Saharan Africa and South Asia with defined catchment areas: Baliakandi and Faridpur, Bangladesh; Kersa, Harar, and Haramaya, Ethiopia; Bamako, Mali; Manhiça and Quelimane, Mozambique; Siaya and Kisumu, Kenya; Makeni and Bo, Sierra Leone; and Soweto, South Africa. Mozambique began enrollment in 2016, followed by South Africa, Mali, Kenya, and Bangladesh in 2017. Enrollment began in 2019 for Ethiopia and Sierra Leone. This analysis used data from stillbirths with complete data by December 31, 2023.

CHAMPS defines stillbirths as fetal deaths without spontaneous breathing or movement at delivery (and no response to resuscitation, if attempted), with a birth weight ≥1000 g or gestational age ≥28 weeks, identified via healthcare facility and community mortality surveillance. Site-specific mortality surveillance and death notification procedures were tailored to local capacity, cultural norms, and geography.^12,13^ Parents or guardians were approached for study enrollment if they lived within the catchment area, and death was reported within 24 hours (72 hours if body was refrigerated).

### Postmortem procedures and data collection

The complete methodology for MITS procedures is described elsewhere.^14–16^ Briefly, trained teams conducted body inspections, anthropometry, and took photographs to assess dysmorphia and maceration. Biopsies of the lungs, brain, and liver, along with available placenta and umbilical cord, were collected using biopsy needles. Tissue underwent histological examination at each site. Samples suggesting infection or diagnostic queries were further evaluated at the US Centers for Disease Control and Prevention. Blood, cerebrospinal fluid (CSF), lung tissue, and rectal swabs were also collected and tested for pathogens using polymerase chain reaction (PCR) on multiplexed TaqMan array cards (Thermo Fisher Scientific, Waltham, MA, USA); blood and CSF underwent convention microbiological culture. Blood samples were also screened for tuberculosis, Human Immunodeficiency Virus (via rapid tests/PCR), and malaria (through thick and thin smears and rapid tests). Clinical data from antenatal maternal health records and hospitalizations for delivery were abstracted, and trained teams interviewed caregivers using the 2016 WHO verbal autopsy instrument. We also explored the relationship between foot length and gestational age, there was a strong relationship between two, but the distributions were much wider to estimate the small for gestational age (SGA) (**Supplementary Figure 1**). However, we classified the stillbirths as small for gestational age (SGA, <10th centile) based on available gestational age, MITS weight, and sex according to the international newborn size for gestational age and sex INTERGROWTH-21st standards.^17^

### Cause of death determination

A detailed description of the determination of cause of death (DeCoDe) process and diagnosis standards are available elsewhere.^18^ In brief, all available data were reviewed by local expert panels (DeCoDe panel) consisting of neonatologists, pediatricians, gynecologists, obstetricians, pathologists, microbiologists, and epidemiologists. Causes of death were recorded using the WHO International Classification of Diseases (ICD-10) and the WHO application of ICD-10 to deaths during the perinatal period (ICD-PM). Conditions in the causal chain were categorized into main and other fetal and maternal conditions, with three confidence levels assigned based on data comprehensiveness and specificity.^15^ For fetal infectious conditions, the DeCoDe panel identified etiologic agents when possible. The panel also evaluated preventable factors, grouping findings for general prevention recommendations. Recognizing the limitations of broad prevention categories, we created more granular recommendation categories and subcategories by analyzing the cases having more detailed descriptions of specific recommendations.

### Statistical Analysis

We used descriptive statistics to summarize categorical variables as frequencies with proportions and continuous variables as medians with interquartile ranges (IQR). Chi-squared test was used to test statistical differences between select groups of interest. Fisher’s exact test was used if the expected value was less than five for more than 20% of cells.

### Ethics Statement

Written consent was obtained for postmortem procedures and data collection, including MITS. Ethics committees in each CHAMPS site and Emory University approved of the study protocols. (https://champshealth.org/resources/protocols/)

## Results

From December 2016 through December 2023, CHAMPS teams approached parents and guardians of 4876 stillbirths eligible for MITS enrollment, 56% (2731/4876) provided consent and had MITS performed, with site-specific consent rates ranging from 32% (432/1356) in Bangladesh to 82% (323/394) in Kenya (**Supplementary Table 1, Supplementary Figure 2**). Cause of death determination was completed for 2492 (91%) enrolled stillbirths and these were included in the analysis; the remainder (n=239) were still pending cause of death determination at the time of analysis. Overall, 51% (1282/2492) of stillbirths with MITS completed had no signs of maceration, and, the proportion with no maceration varied significantly across sites, ranging from 27% (84/306) in South Africa to 61% (303/498) in Mozambique (**Table 1**). Sixty-three percent (1577/2492) weighed <2,500 g at the time of MITS, including 27% (671/2492) weighing <1500 g. Twenty-one percent of stillbirths had an unknown gestational age, precluding our ability to discern if these babies were small-for-gestational age. However, among those (1726/2492) with information available on gestational age and MITS weight, 36% (625/1726) were small for gestational age, again with high variation across sites, ranging from 11% (61/568) in Ethiopia to 45% (184/405) in Bangladesh. Seventy-five percent (1878/2492) were vaginal deliveries and 95% (2361/2492) were delivered at a healthcare facility (**Table 1**). No documentation on the presence of fetal heartbeat during labour was available in over 50% of cases (**Table 1**). The MITS procedure was conducted within 24 hours of birth for 91% (2264/2492) of stillbirths. (**Table 1**). Thirty-two maternal deaths were reported in clinical records of these stillbirths, although no information about the cause of death was available for these women.

**Table 1.**
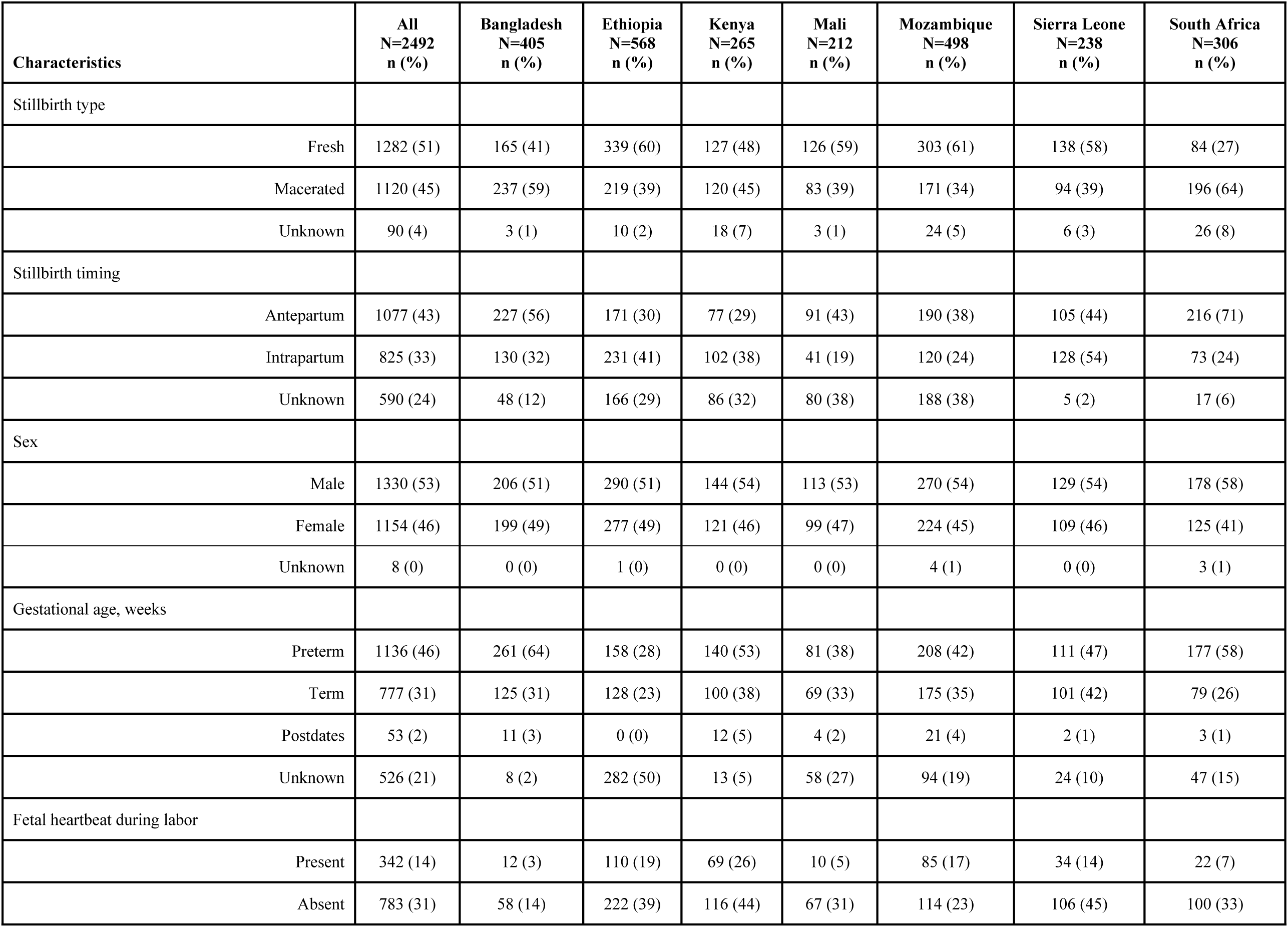
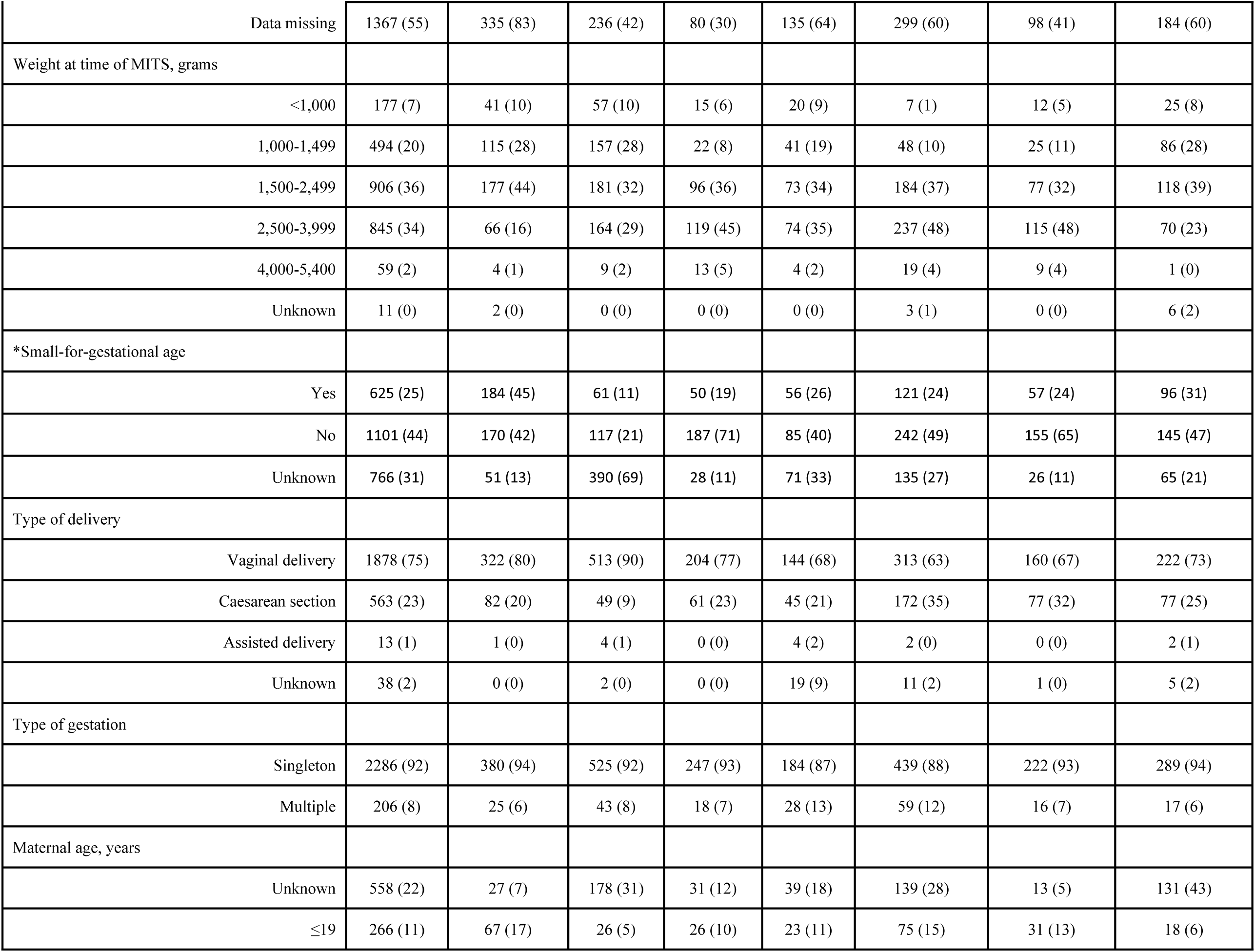
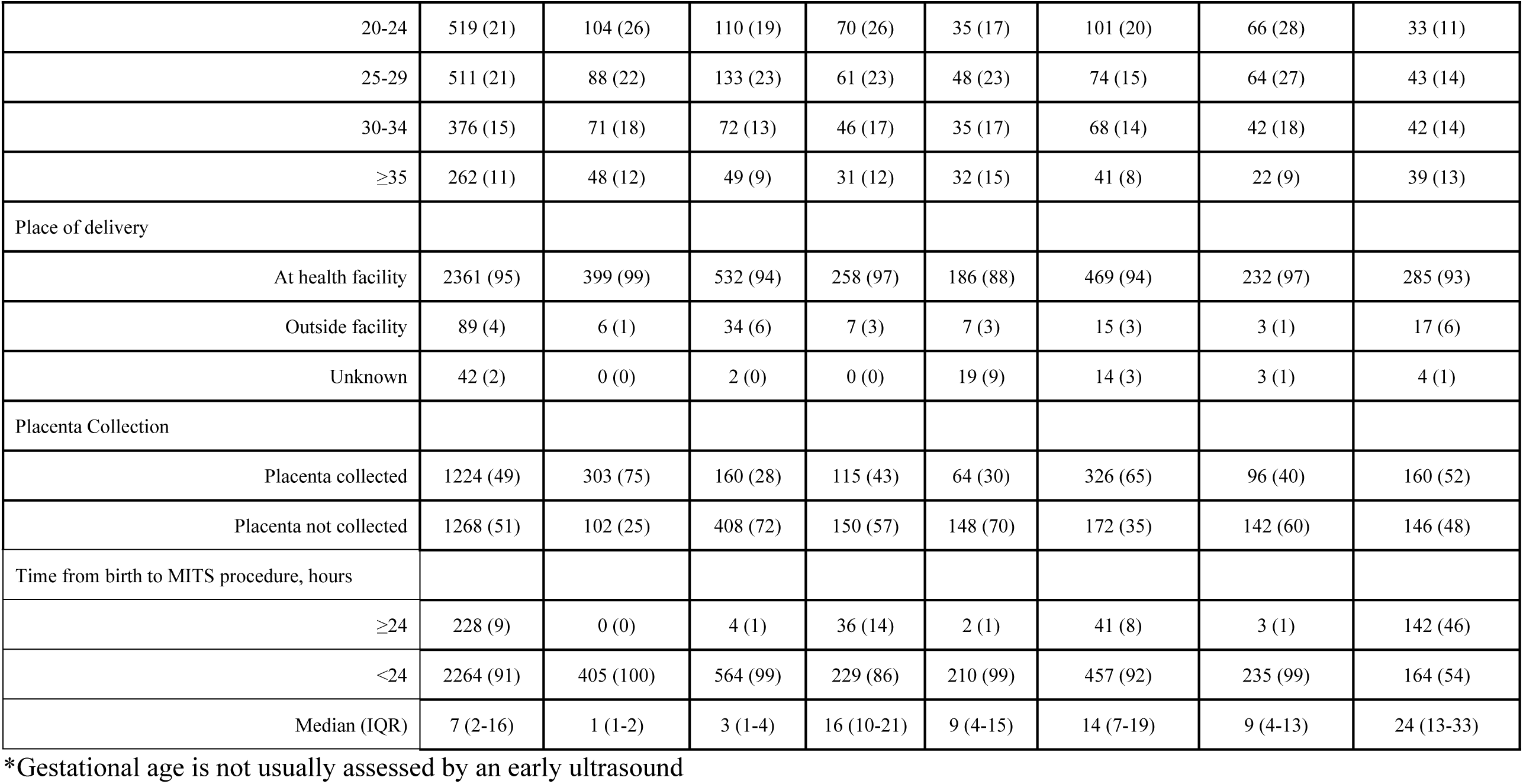
Characteristics of 2492 stillbirths with cause of death determined, by CHAMPS site, 2016-2023.

### Conditions in the fetus leading to death

DeCoDe panels were able to determine a cause of death (fetal or maternal condition) for 94% (2342/2492) of stillbirths reviewed. Among those with a cause determined, at least one main condition in the fetus leading to death was identified in 97% (2278/2342) (**Supplementary Table 2)**; 64 stillbirths (3%) had other fetal conditions in the causal chain. DeCoDe panels noted the strongest level of confidence (level 1) for the evidence supporting the main condition in the fetus for 81% (1889/2492) (**Supplementary Table 3**). Intrauterine hypoxia, defined as the hypoxic condition of a fetus starting before or during labor, was determined to be the main condition in the fetus in 75% (1864/2492) of all stillbirths and was consistently the predominant main condition regardless of site, ranging from 59% (182/306) in South Africa to 89% (236/265) in Kenya (**Figure 1, Supplementary Table 2**). For 18% (322/1864) of the cases with intrauterine hypoxia as the main condition, no underlying maternal or placental causes were identified (**Supplementary Table 4**). Congenital birth defects accounted for 9% (218/2492) of main conditions in the fetus, ranging from 2% (4/265) in Kenya to 24% (134/568) in Ethiopia (**Figure 1, Supplementary Table 2**). Sixty-two percent (135/218) of congenital birth defects were neural tube defects, most of which (82%, 110/134) occurred in the Ethiopia site. Congenital infection was the third most common condition in the fetus for 8% (200/2492) of all stillbirths, ranging from 1% (3/238) in Sierra Leone to 28% (87/306) in South Africa (**Supplementary Table 2**).

**Figure 1.**
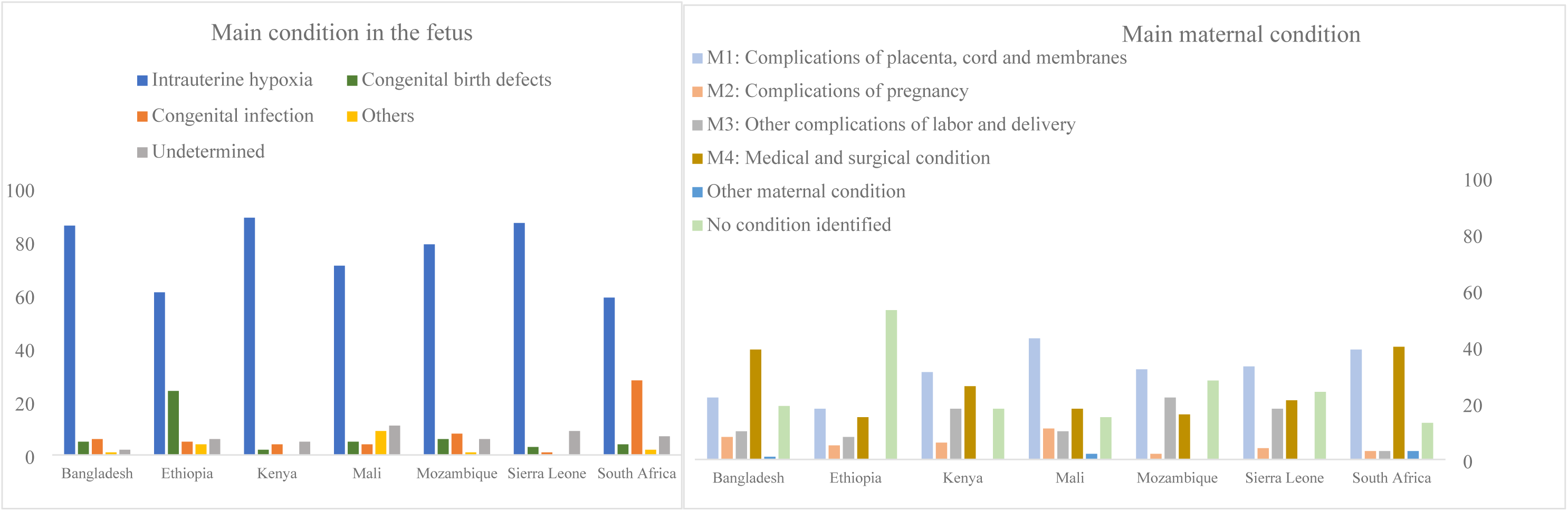
Causes of death among 2492 stillbirth cases with complete cause of death determination, by CHAMPS site, 2016-2023.

### Etiologies of congenital infections

A total of 245 etiologic agents were identified in the causal chain among the 228 stillbirths with congenital infection (**Supplementary Table 2)**. Sixty-six percent had one etiologic agent detected (150/228), 12% (27/228) had two, 5% (11/228) had three, and 1% (2/228) had four. Forty-five percent of all stillbirths with congenital infection in the causal chain had one or more gram-positive bacteria (102/228), 51% (116/228) had one or more gram-negative bacteria, 10% (23/228) had one virus, 1% (2/228) had one protozoa and 1% (1/228) had one fungus (**Supplementary Table 2, Supplementary Table 5**).

The primary infectious causes of stillbirths were Group B Streptococcus (2%, 46/2492), *Escherichia coli* (2% 43/2492), and *Enterococcus faecalis* (1%, 24/2492) (**Supplementary Table 5**). Almost all viral infections were due to cytomegalovirus, contributing to 1% of all stillbirths (20/2492). Congenital syphilis was only reported in South Africa (5%, 14/306), Mozambique (1%, 4/498), and Bangladesh (0.5%, 2/405); **Supplementary Table 5)**.

### Comparison of conditions in antepartum and intrapartum stillbirths

Seventy-six percent (1902/2492) of stillbirths had information available to determine whether the death occurred antepartum or intrapartum (**Table 1, Supplementary Table 6**). The distribution of the main condition in the fetus across the antepartum, intrapartum, and stillbirths with unknown timing were similar (**Supplementary Table 7**). Most stillbirths with an undetermined condition in the fetus were antepartum stillbirths (53%, 79/150). Macerated stillbirths accounting for a higher proportion of antepartum cases compared to stillbirths without maceration (67%, 724/1077 vs 30%, 322/1077) (**Supplementary Table 7)**.

### Main maternal condition contributing to fetal death

We identified a maternal condition contributing to fetal death in 72% (1794/2492) of stillbirths. In 5% (122/2492) of cases, maternal conditions remained unknown due to lack of maternal data collected. Additionally, in 23% (576/2492), specific maternal conditions could not be identified even though maternal medical records were available; poor quality and completeness of records contributed to many of these cases (**Supplementary Table 8**). The most common group of maternal conditions based on ICD-PM classifications was complications of the placenta, cord, and membranes (29%, 724/2492) (**Figure 1, Supplementary Table 2**). The second most common group was medical and surgical conditions (24%, 602/2492) (**Figure 1, Supplementary Table 2**), largely consisting of maternal hypertension (17%, 414/2492) (**Supplementary Table 4**).

Forty-nine percent (1224/2492) of stillbirths had placenta collected, with significant variation across sites, ranging from 28% (160/568) in Ethiopia to 75% (303/405) in Bangladesh (**Table 1)**, and 33% (409/1224) of stillbirths with a placenta examined were attributed to placental and membrane related complications (**Supplementary Table 3**). There was no correlation between the collection and investigation of the placenta for stillbirths and the proportion of stillbirths where a main maternal condition was identified across sites. (**Supplementary Figure 2**) Among the 1268 stillbirths where placenta was not available, placenta-related complications were determined based only on maternal clinical records and verbal autopsy interviews and were less likely to be attributed to placental and membrane cord complications than those with a placenta available for investigation (17%, 210/1268) (**Supplementary Table 3**).

Placental complications (22%; 415/1864) and maternal hypertension (20%; 382/1864) were the primary conditions among the subset of stillbirths with intrauterine hypoxia as the main condition in the fetus (**Supplementary Table 4).** Chorioamnionitis and membrane complications (36%, 73/200) and maternal infection (12%, 25/200) were the primary maternal conditions among stillbirths with congenital infections. We identified only 9 stillbirths where the maternal infectious cause was HIV infection (**Supplementary Table 4**), reported from three countries: Kenya (5/265), Sierra Leone (2/238), and South Africa (2/306). No maternal condition was identified for 88% (191/218) of stillbirths with congenital birth defects as the main fetal condition. Among stillbirths classified as small-for-gestational-age (36%, 625/1726), the most frequently identified contributing conditions were maternal hypertension (23%, 141/625) and placental complications (21%, 129/625; **Supplementary Table 9**).

### Prevention

Seventy-two percent (1808/2492) of stillbirths were considered preventable by DeCoDe panels, under optimal provision of existing care (**Table 2**). Improving antenatal care and obstetric care and management could have prevented 85% of stillbirths (1532/1808) and was the most frequently recommended intervention across all sites. A further 27% could have been prevented by improved health-seeking behavior on the part of families (491/1808) with the highest proportion reported in Ethiopia (57%, 267/470). Recommendations also emphasized improving family planning in Ethiopia (53%, 247/470) and prioritized HIV prevention and control in Kenya (24%, 60/250) (**Supplementary Table 10)**. Among 1099 (44%) stillbirths with more details about recommended prevention measures, 69% (753/1099) could have been prevented through improved antenatal care, which includes quality antenatal care (31%, 338/1099), management of medical conditions (30%, 328/1099) and health education (24%, 268/1099) as the three most common subcategories (**Figure 2)**. Thirty-three percent of stillbirths could have been prevented through improved obstetric management (361/1099) and 10% through delivery by a qualified provider at a healthcare facility (105/1099); 9% could have been saved through improved documentation of medical records (98/1099) (**Figure 2)**. Forty percent of congenital infections were not considered preventable (**Table 2**), primarily because there are currently no maternal screening guidelines or interventions for the causative pathogens.

**Table 2.**
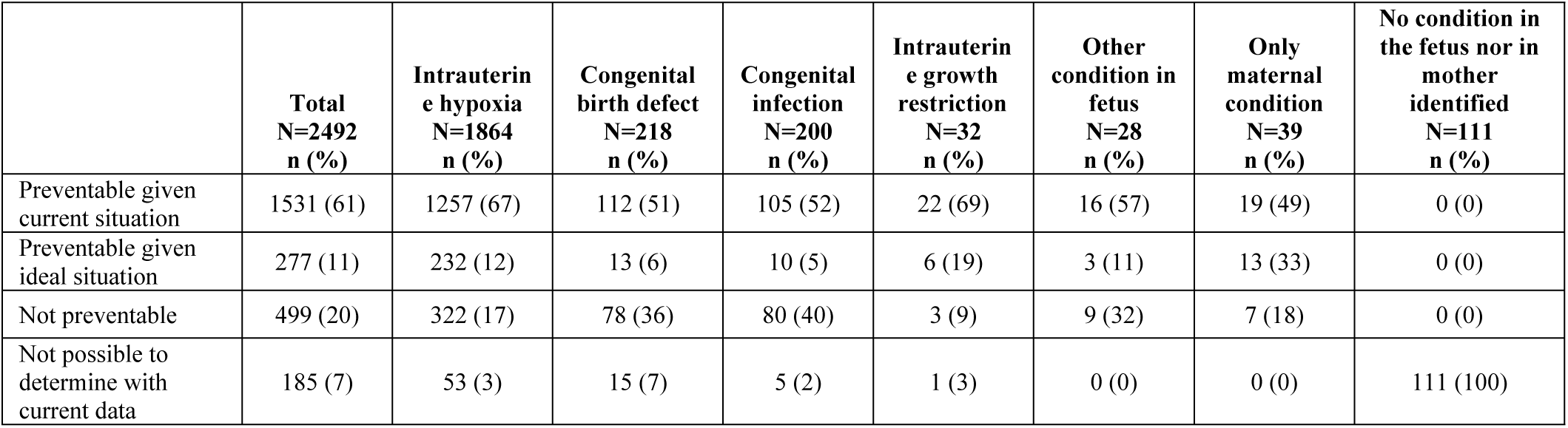
Number of preventable stillbirths with complete cause of death determination, by main condition in the fetus, 2016-2023.

|  | **Total**<br>**N=2492**<br>**n (%)** | **Intrauterin**<br>**e hypoxia**<br>**N=1864**<br>**n (%)** | **Congenital**<br>**birth defect**<br>**N=218**<br>**n (%)** | **Congenital**<br>**infection**<br>**N=200**<br>**n (%)** | **Intrauterin**<br>**e growth**<br>**restriction**<br>**N=32**<br>**n (%)** | **Other**<br>**condition in**<br>**fetus**<br>**N=28**<br>**n (%)** | **Only**<br>**maternal**<br>**condition**<br>**N=39**<br>**n (%)** | **No condition in**<br>**the fetus nor in**<br>**mother**<br>**identified**<br>**N=111**<br>**n (%)** |
| --- | --- | --- | --- | --- | --- | --- | --- | --- |
| Preventable given current situation | 1531 (61) | 1257 (67) | 112 (51) | 105 (52) | 22 (69) | 16 (57) | 19 (49) | 0 (0) |
| Preventable given ideal situation | 277 (11) | 232 (12) | 13 (6) | 10 (5) | 6 (19) | 3 (11) | 13 (33) | 0 (0) |
| Not preventable | 499 (20) | 322 (17) | 78 (36) | 80 (40) | 3 (9) | 9 (32) | 7 (18) | 0 (0) |
| Not possible to determine with current data | 185 (7) | 53 (3) | 15 (7) | 5 (2) | 1 (3) | 0 (0) | 0 (0) | 111 (100) |

**Figure 2:**
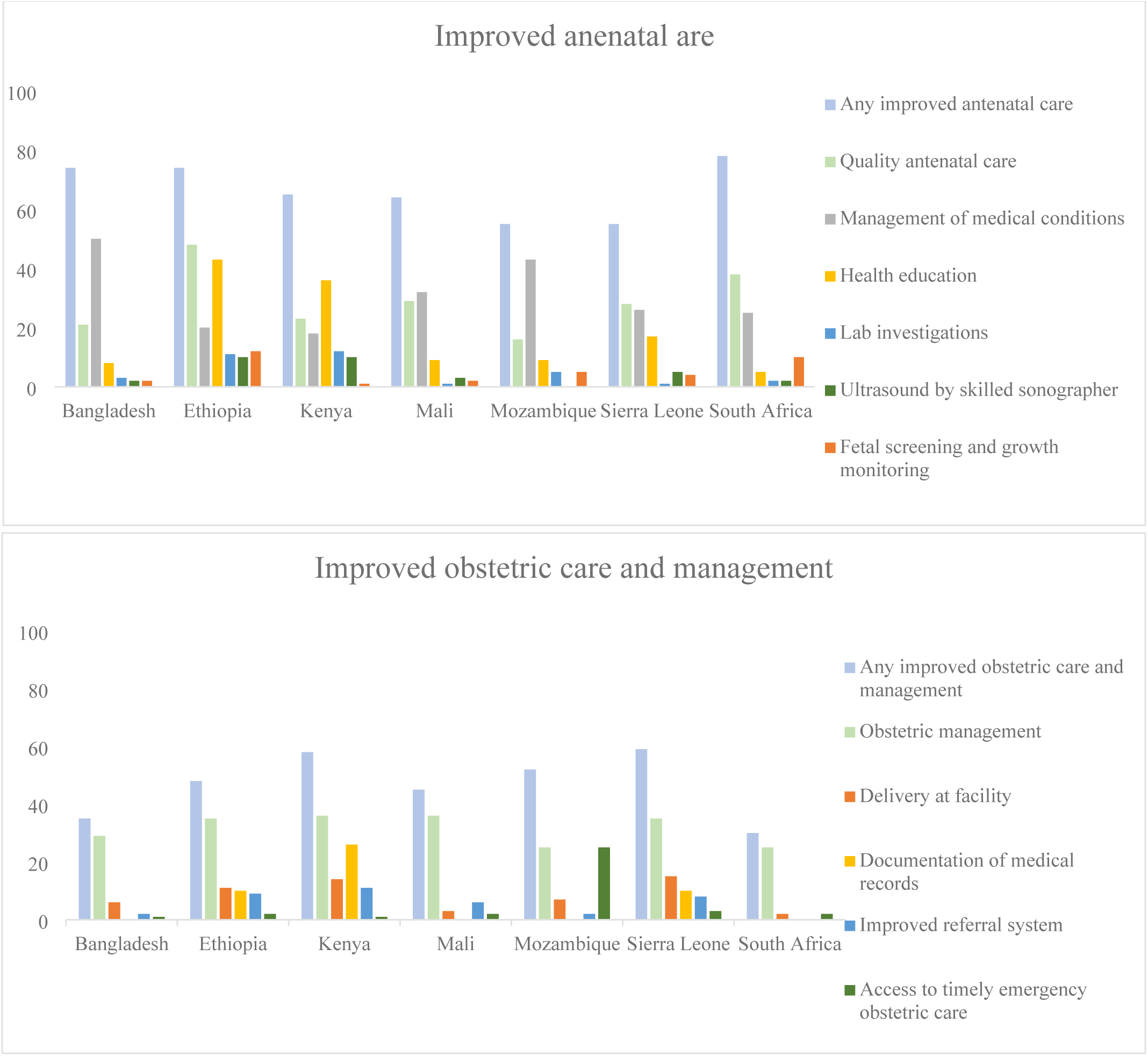
Detailed recommendations regarding antenatal care and obstetric care and management, 1099 preventable stillbirth cases with recommendation descriptions, by CHAMPS site, 2016-2023.

## Discussion

Our investigation of 2492 stillbirths across seven years in seven countries identified intrauterine hypoxia linked to maternal conditions like hypertension or placenta or cord abnormalities as the most common causes, with congenital infections, and birth defects as leading causes when a condition was identified in the fetus. Our findings support earlier findings from CHAMPS and the PURPOSe study. However, the scope and scale of our study allowed us to identify important geographic differences in the causes of stillbirths across countries, likely due to heterogeneity in access to quality antenatal and obstetric care. For example, in Mozambique, Mali, and Bangladesh, the main strategy for preventing stillbirths during antenatal care was managing maternal medical conditions. In contrast, in Ethiopia, Sierra Leone, and South Africa, improving the quality of antenatal care was more important. In Mozambique, access to timely and appropriate obstetric care was a major gap, which was less of an issue in other countries. These differences highlight the need to tailor prevention strategies to the specific healthcare context of each country. For instance, folate fortification in Ethiopia will be key to reducing neural tube defects, while this cause of death is uncommon in other sites, all of which have folate-fortified foods.^19^ Similarly, congenital infections were a significant contributor to stillbirths in South Africa compared to other locations.

Approximately three-quarters of stillbirths had intrauterine hypoxia as the main fetal condition. This diagnosis implies that no specific abnormality or disease was identified in the fetus, but rather suggests that pregnancy or delivery complications led to stillbirth. The two most common maternal conditions identified among stillbirths with intrauterine hypoxia were maternal hypertension and placental complications, highlighting missed opportunities for prevention from preconception to antenatal care and during labor and delivery by improved identification, monitoring, and treatment. Over the past two decades, pregnancy-related hypertensive disorders and broadly adult hypertension have remained highly prevalent in sub-Saharan Africa and South Asia.^20,21^ Globally, the proportion of pregnancies affected by hypertensive disorders increased by 11% from 1990 to 2019.^21^ Nevertheless, pregnant women experience a series of barriers in managing hypertension and, ultimately, in preventing stillbirths. A large proportion of individuals (40.5%) were unaware of their hypertensive status until screened^22^, which could promote awareness of chronic hypertension before conception, and inadequate knowledge of hypertensive disorders in pregnancy-41.8% of pregnant women in Ethiopia lacked awareness of pregnancy-induced hypertension.^23^ These challenges are compounded by inadequate access or sociocultural barriers to antenatal care,^24^ poor screening practices when antenatal care is sought (only 31% received WHO-recommended screening),^25,26^ and poor management of hypertensive disorders when detected (Among 25 cases, only 15/25 women diagnosed with severe PE/E received MgSO4, three of those cases were treated with both MgSO4 and diazepam, a potentially dangerous practice which is not based on global recommendations. In addition, 13 women received anti-hypertensives while five did not).^25,26^ Other complications, such as placental abruptions, can be challenging to prevent with current medical technologies but have established clinical guidelines related to intrapartum care based on the level of maternal and fetal compromise and also gestational age.^27^

Systematic protocols are in place across CHAMPS sites for DeCoDe panel review, however, we recognized some heterogeneity in fetal and maternal or placental conditions determined to be the cause of stillbirths. This variation may be due to site-to-site differences in how stillbirths are identified and enrolled, the availability of maternal medical records and placental examinations, and the relative familiarity of each site’s expert panel with specific causes. We identified important geographic heterogeneity in stillbirth from congenital infections; nearly a third of all stillbirths in Soweto, South Africa, were attributed to congenital infections compared to 2-10% at other sites. The most common pathogens causing congenital infections, including Group B Streptococcus, *Escherichia coli*, and Enterococcus faecalis, are pathogens commonly associated with rectovaginal tract colonization in pregnant women, as well as invasive infections among stillbirths in Soweto.^28,29^ Although these pathogens were found at other sites, they accounted for only a small proportion of stillbirths relative to those linked to pregnancy and delivery complications at other sites. These congenital infections may be higher in South Africa because the quality of antenatal and perinatal care is better there, reducing the burden from those care gaps. This variation in cause-specific mortality suggests that targeted interventions, such as maternal GBS vaccines^28^, could be more beneficial in South Africa, whereas better access to quality prenatal and delivery care is urgently needed in other CHAMPS sites. We also observed heterogeneity in congenital birth defects, which contributed to nearly a quarter of stillbirths in Eastern Ethiopia. Approximately half of all congenital birth defects were neural tube defects primarily identified in Ethiopia, a condition that can largely be prevented through folic acid supplementation.^30^ Recent findings from CHAMPS suggest countries with mandatory folic acid fortification and high fortification coverage are successfully preventing neural tube defects.^19^

Among the stillbirths where we had information about gestational age, over one-third were small for gestational age (SGA). Previous studies have classified as small-for-gestational-age stillbirths as part of the small vulnerable newborns group, but they have not been fully integrated into burden assessments. In our study, the majority of SGA-related stillbirths were attributed to maternal hypertension and placental complications. With better antenatal care, these small vulnerable stillbirths could be identified early in pregnancy, which is a potential area of intervention to prevent SGA-related stillbirths.

The main limitation of our study is that incomplete maternal records, including missing gestational age (21%), timing of fetal death (24%) and discarded placentas (51%), meant that key pieces of information were unavailable for a large proportion of stillbirths. Placenta collection can be challenging as the placenta is not routinely or properly preserved at health facilities, study consent often occurs hours after delivery, when the placenta may have already been discarded, and many stillbirths occur outside of health facilities, making the retrieval of the placenta difficult. Placental specimens were collected from half of all stillbirths enrolled in CHAMPS; enhanced efforts to obtain and investigate placentas would improve the ability to attribute stillbirths to placental causes. The placentas collected were investigated for gross and histological examination; however, some infectious causes may have gone undetected due to the lack of testing of placentas for pathogens. The PURPOSe study, for example, identified *Ureaplasma urealyticum* in 39% of placentas they studied using molecular tests. Our current understanding of some maternal conditions is primarily based on maternal medical records and verbal autopsy, which were frequently unavailable or incomplete, either because tests were not conducted or the results were not recorded. Incomplete maternal records, including lack of fetal heartbeat monitoring during delivery, likely limited our ability to identify causes of stillbirths and suggest prevention strategies. Often, places with the least amount of data available are those with the highest rates of stillbirths, compounding the problem of prevention. A comprehensive investigation into the mother’s health, including recommended laboratory testing and fetal monitoring, would enhance our understanding of the causal chain and opportunities for prevention, and would also likely improve outcomes, as these form part of quality antenatal and obstetric care. Performing routine karyotyping or, preferably, chromosomal microarray analysis on the stillbirths in this study would likely have influenced the distribution of proportions assigned to various causes especially valuable in analyses of stillbirths with congenital anomalies.^32^ Lastly, despite significant training and efforts to standardize DeCoDe panels across our sites, some variation in cause of death determination and recommended prevention strategies across sites may be due to differences in interpretation of data. For example, the importance of health education as a preventive measure was frequently identified in Kenya and Ethiopia, though it was less emphasized in other sites.

CHAMPS is the longest-running systematic postmortem investigation into causes of stillbirths, and our findings are likely representative of stillbirths occurring at each site.^19^ This analysis solidifies our prior understanding of the causes of stillbirths and identifies some geographic heterogeneity in the proportion of stillbirths linked to major causes. Furthermore, it highlights the myriad persistent gaps in access to quality maternal and obstetric care, and subsequent gaps in information about the stillbirth, that must be urgently addressed to reduce stillbirths and better define their underlying causes.

### CHAMPS Consortium

For CHAMPS Bangladesh: ASM Nawshad Uddin Ahmed and Mahbubul Hoque of Dhaka Shishu Hospital and Institute; Mohammed Kamal, Mohammad Mosiur, and Ferdousi Begum of Bangabandhu Sheikh Mujib Medical University; Saria Tasnim of Dhaka Community Medical College and Hospital; Meerjady Sabrina Flora of the Directorate General of Health Services in Bangladesh; Farida Arjuman of Institute of Cancer Research and Hospital, Tahmina Shirin, and Mahbubur Rahman of Institute of Epidemiology, Disease Control and Research; Sanwarul Bari, Shahana Parveen, Mustafizur Rahman, Dilruba Ahmed, of International Centre for Diarrhoeal Disease Research, Bangladesh; and Ferdousi Islam of Popular Medical College and Hospital in Dhaka, Bangladesh. For CHAMPS Ethiopia: Joseph O Oundo of London School of Hygiene & Tropical Medicine; Fikremelekot Temesgen of Addis Ababa University in Addis Ababa, Ethiopia; Melisachew Mulatu Yeshi of Mekelle University, Mekele, Ethiopia; Alexander M Ibrahim, Tadesse Gure, Addisu Alemu of the College of Health and Medical Sciences at Haramaya University, Stian MS Orlien of the University of Hargeisa, Somaliland and Vestfold Hospital Trust, Tønsberg, Norway; and Mahlet Abayneh Gizaw of St Paul’s Hospital Millennium Medical College in Addis Ababa, Ethiopia. For CHAMPS Kenya: Aggrey Igunza, Peter Otieno, Janet Agaya, Richard Oliec, Joyce Akinyi Were, Dickson Gethi, Sammy Khagayi, George Aol, Thomas Misore, Harun Owuor, Christopher Muga, Bernard Oluoch, Christine Ochola, Peter Nyamthimba of Kenya Medical Research Institute. For CHAMPS Mali: Karen D Fairchild of University of Virginia; Carol L Greene, Rima Koka, Sharon M Tennant, Ashka Mehta, Brigitte Gaume, Sharon M. Tennant, Carol L. Greene, Kiranpreet Chawla, and J Kristie Johnson of University of Maryland School of Medicine; Adama Mamby Keita, Nana Kourouma, Uma U Onwuchekwa, Awa Traore, Jane Juma, Doh Sanogo, Diakaridia Sidibe, Cheick Bougadari Traore, Kounandji Diarra, Tiéman Diarra, and Seydou Sissoko of Centre pour le Développement des Vaccins; and Diakaridia Kone of CSRef Commune I in Bamako, Mali. For CHAMPS Mozambique: Khátia Munguambe, Ariel Nhacolo, Tacilta Nhampossa of Centro de Investigação em Saúde de Manhiça in Maputo; Maria Maixenchs, Clara Menéndez of IS Global Hospital Clinic at Universität de Barcelona; Zara Manhique of Quelimane Central Hospital; and Sibone Mocumbi of University and Maputo Central Hospital. From the CHAMPS Program Office: Jeffrey P Koplan, Mischka Garel, Kurt Vyas, Courtney Bursuc, Valentine Wanga, Kristin LaHatte, Sarah Raymer, John Blevins, Solveig Argeseanu, Margaret Basket and Manu Bhandari of Emory Global Health Institute; and Shailesh Nair, Navit T Salzberg, and Lucy Liu of the Public Health Informatics Institute at the Task Force for Global Health in Atlanta, GA, USA. From the Centers for Disease Control and Prevention Central Pathology Lab: Jana M Ritter, Ashutosh Wadhwa and Tais Wilson of Infectious Diseases Pathology Branch, National Center for Emerging and Zoonotic Diseases at the Centers for Disease Control and Prevention. For the Centers for Disease Control and Prevention TaqMan Array Team: Jonas M Winchell, Jacob Witherbee, and Jessica L Waller of the National Center for Immunization and Respiratory Diseases at the Centers for Disease Control and Prevention. For the Centers for Disease Control and Prevention: Roosecelis Martines, Shamta Warang, Maureen Diaz, Jessica Waller, Zachary Madewell. For CHAMPS Sierra Leone: Dickens Kowuor, Andrew Moseray, Julius Ojulong, Erick Kaluma, Soter Ameh, and Oluseyi Balogun of Crown Agents; Samuel Pratt of FOCUS 1000; and Carrie-Jo Cain and Solomon Samura of World Hope International. Fatmata Bintu Tarawally, Martin Seppeh, Ronald Mash, Babatunde Duduyemi, James Bunn (WHO Int), Alim Swaray-Deen, Joseph Bangura, Amara Jambai, Margaret Mannah, Okokon Ita, Cornell Chukwuegbo, Sulaiman Sannoh, Princewill Nwajiobi, Francis Moses, Tom Sesay, James Squire, Joseph Kamanda Sesay, Osman Kaykay, Binyam Halu, Hailemariam Legesse (unicef), Francis Smart, Sartie Kenneh, Soter Ameh, Sartie Kenneh. For CHAMPS South Africa: Fatima Solomon, Megan Dempster, Siobhan Johnstone, Takwanisa Machemedze, Marguerite Hall, Shabnam Shaik, Bongani Ntimani, Sarah Downs, Avani Kashiram, Michelle Groome, Alane Izu, Yasmin Adam of South African Council Vaccines and Infectious Diseases Analytics Research Unit, University of Witwatersrand; Sithembiso Velaphi, Sanjay Lala, Martine Hale, Firdose Nakwa, Tanya Ruder, Prenika Jaglal of Chris Hani Baragwanath Academic Hospital and University of Witwatersrand; Lesego Mothibi of National Health for Laboratory Service in South Africa; Amy Wise of Wits Department of Obstetrics & Gynaecology, Faculty of Health Sciences, University of the Witwatersrand, Johannesburg, South Africa; Vuyelwa Baba, Philiswa Mlandu, Washington Mudini.

### Contributors

RFB designed the project and acquired the grant funds. AR, ESG, and KL developed the protocol and conducted the analysis. DMB and KL directed the data management. AR, ESG, KL, CGW, JAGS drafted and revised the manuscript. SEA, AIC, MA, AA, IO, SS, EK, SS, AMK, MT, AM, KK, SAM, SM, YA, AW, ZD, CM, SS, EO, RO, AKI, DO, VA, IM, QB, SA, RV, EX, NA, LM, MMY, FT, LL, BD, and PM participated in protocol development and coordinated the clinical and diagnostic data collection. The final decision to submit the manuscript for publication was made by AR, ESG, CGW, and KL. The manuscript received a thorough review and unanimous approval from all authors for submission.

## Declaration of interests

CGW reports other grants from the Bill & Melinda Gates Foundation for work on malnutrition, HIV, and Covid vaccines, other than the submitted work. AW reports being a council member of the national Obstetrician and & Gynaecologists council. KK reports grant from the Bill & Melinda Gates Foundation and support for attending meetings. SEA reports his employment at icddr,b, an organization that receives grants for related projects and funding from WHO & BMGF to support his travels. All other authors declare no competing interests.

## Data sharing

CHAMPS data are available online and requests for further detailed data for research and evaluation purposes can be made at: https://champshealth.org/data/.

## Acknowledgements

This project received funding from the Bill & Melinda Gates Foundation (Grant No. OPP1126780). The CHAMPS network extends earnest gratitude to the families who took part in this study. The network also acknowledges the efforts of individuals involved in minimally invasive tissue sampling (MITS), the demographic surveillance system, social behavioral science, informatics, and laboratory activities, along with the local communities at each of the seven sites. Several authors are affiliated with the US Centers for Disease Control and Prevention (CDC). However, the findings and conclusions presented in this document are solely those of the authors and do not necessarily represent the official position of the US Centers for Disease Control and Prevention.

## Supplementary appendix

Supplement to: Rahman A, Lee K, Arifeen SE, et al. Causes of stillbirth in sub-Saharan Africa and South Asia: Findings from the Child Health and Mortality Prevention Surveillance Network, 2016-2021.

**Supplementary Figure 1.**
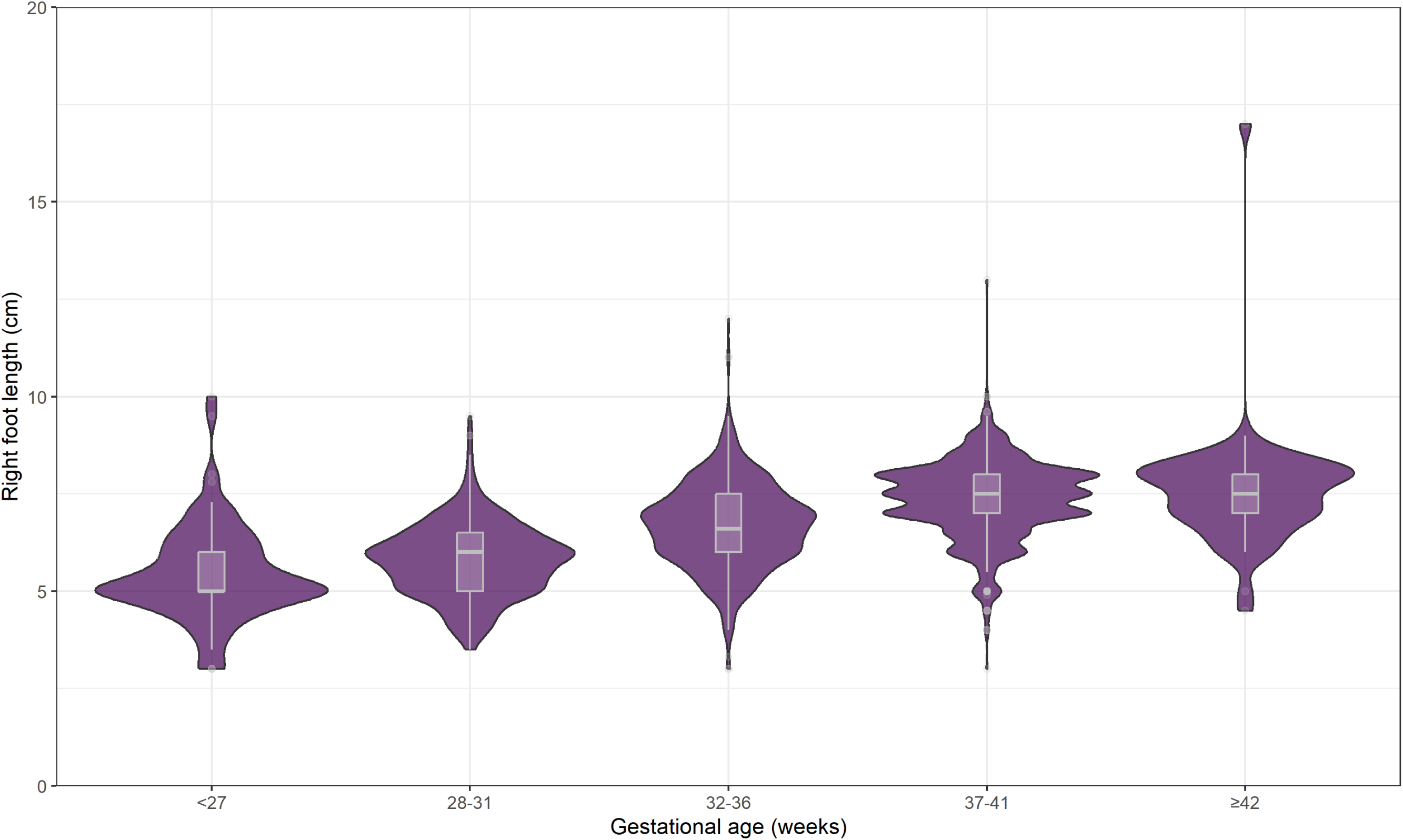
Relationship between foot length and gestational age among 1966 stillbirths enrolled in minimally invasive tissue sampling and known gestational age (N=2492), 2016-2023.

**Supplementary Table 1.** Number of stillbirths eligible for enrollment in CHAMPS, underwent minimal invasive tissue sampling, completed cause of death determination, and ascertained cause of death, by CHAMPS site, 2016-2023.

|  | **All eligible for**<br>**enrollment**<br>**N=4876**<br>**n (%)** | **Bangladesh**<br>**N=1356**<br>**n (%)** | **Ethiopia**<br>**N=935**<br>**n (%)** | **Kenya**<br>**N=394**<br>**n (%)** | **Mali**<br>**N=635**<br>**n (%)** | **Mozambique**<br>**N=719**<br>**n (%)** | **Sierra Leone**<br>**N=347**<br>**n (%)** | **South Africa**<br>**N=490**<br>**n (%)** |
| --- | --- | --- | --- | --- | --- | --- | --- | --- |
| Underwent minimally invasive tissue sampling | 2731 (56) | 432 (32) | 608 (65) | 323 (82) | 274 (43) | 507 (71) | 279 (80) | 308 (63) |
| Completed cause of death determination | 2492 (51) | 405 (30) | 568 (61) | 265 (67) | 212 (33) | 498 (69) | 238 (69) | 306 (62) |
| Ascertained cause of death | 2381 (49) | 396 (29) | 537 (57) | 262 (66) | 202 (32) | 470 (65) | 222 (64) | 292 (60) |

**Supplementary Figure 2.**
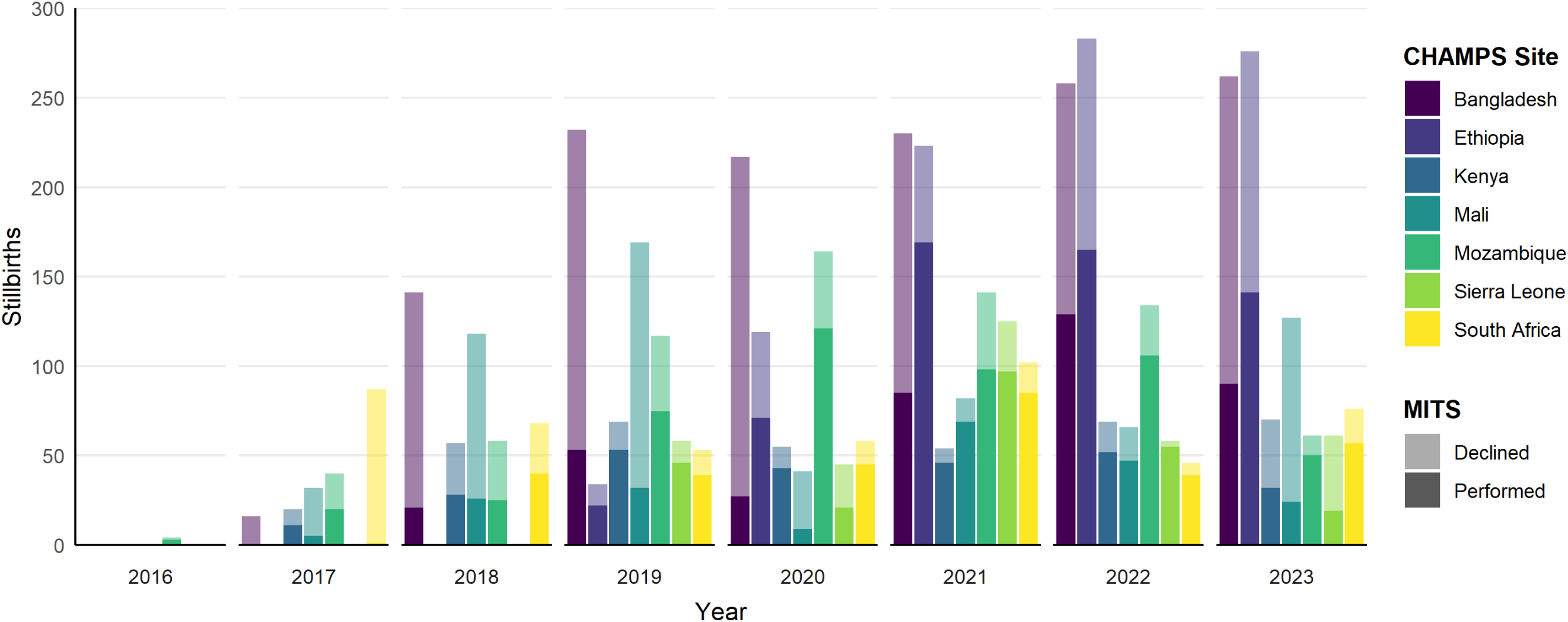
Number of stillbirths eligible for minimally invasive tissue sampling (N=2492), by year, site, and consent status.

**Supplementary Table 2.** Causes of death among 2492 stillbirth cases with complete cause of death determination, by CHAMPS site, 2016-2023.

| Cause of death | All<br>N=2492<br>n (%) | Bangladesh<br>N=405<br>n (%) | Ethiopia<br>N=568<br>n (%) | Kenya<br>N=265<br>n (%) | Mali<br>N=212<br>n (%) | Mozambique<br>N=498<br>n (%) | Sierra Leone<br>N=238<br>n (%) | South Africa<br>N=306<br>n (%) |
| --- | --- | --- | --- | --- | --- | --- | --- | --- |
| Main condition in the fetus |  |  |  |  |  |  |  |  |
| Intrauterine hypoxia | 1864 (75) | 347 (86) | 346 (61) | 236 (89) | 151 (71) | 395 (79) | 207 (87) | 182 (59) |
| Congenital birth defects | 218 (9) | 21 (5) | 134 (24) | 4 (2) | 11 (5) | 28 (6) | 7 (3) | 13 (4) |
| Congenital infection | 200 (8) | 24 (6) | 29 (5) | 10 (4) | 8 (4) | 39 (8) | 3 (1) | 87 (28) |
| Others | 60 (2) | 4 (1) | 24 (4) | 2 (0) | 19 (9) | 7 (1) | 0 (0) | 4 (2) |
| Undetermined | 150 (6) | 9 (2) | 35 (6) | 13 (5) | 23 (11) | 29 (6) | 21 (9) | 20 (7) |
| Other condition in the fetus<sup>1</sup> |  |  |  |  |  |  |  |  |
| Intrauterine hypoxia | 113 (5) | 8 (2) | 74 (13) | 0 (0) | 20 (9) | 3 (1) | 5 (2) | 3 (1) |
| Intrauterine growth restriction | 53 (2) | 51 (13) | 0 (0) | 1 (0) | 0 (0) | 0 (0) | 0 (0) | 1 (0) |
| Congenital infection | 28 (1) | 2 (0) | 14 (2) | 0 (0) | 1 (0) | 0 (0) | 2 (1) | 9 (3) |
| Preterm birth complications | 25 (1) | 11 (3) | 2 (0) | 0 (0) | 0 (0) | 2 (0) | 10 (4) | 0 (0) |
| Congenital birth defects | 9 (0) | 6 (1) | 2 (0) | 0 (0) | 0 (0) | 1 (0) | 0 (0) | 0 (0) |
| Other | 6 (0) | 2 (0) | 1 (0) | 0 (0) | 0 (0) | 0 (0) | 2 (1) | 1 (0) |
| Aspiration syndrome | 2 (0) | 2 (0) | 0 (0) | 0 (0) | 0 (0) | 0 (0) | 0 (0) | 0 (0) |
| Birth trauma | 1 (0) | 0 (0) | 0 (0) | 0 (0) | 0 (0) | 0 (0) | 1 (0) | 0 (0) |
| Main maternal condition (ICD-PM) |  |  |  |  |  |  |  |  |
| M1: Complications of placenta, cord and membranes | 724 (29) | 91 (22) | 102 (18) | 83 (31) | 92 (43) | 157 (32) | 79 (33) | 120 (39) |
| M2: Complications of pregnancy | 131 (5) | 32 (8) | 30 (5) | 15 (6) | 24 (11) | 12 (2) | 10 (4) | 8 (3) |
| M3: Other complications of labor and delivery | 319 (13) | 42 (10) | 47 (8) | 47 (18) | 21 (10) | 109 (22) | 44 (18) | 9 (3) |
| M4: Medical and surgical condition | 602 (24) | 158 (39) | 86 (15) | 70 (26) | 39 (18) | 79 (16) | 49 (21) | 121 (40) |
| Other maternal condition | 18 (1) | 4 (1) | 1 (0) | 1 (0) | 4 (2) | 0 (0) | 0 (0) | 8 (3) |
| No condition identified | 698 (28) | 78 (19) | 302 (53) | 49 (18) | 32 (15) | 141 (28) | 56 (24) | 40 (13) |
| Etiologic agents |  |  |  |  |  |  |  |  |
| Gram-positive bacteria | 102 (4) | 8 (2) | 18 (3) | 4 (2) | 5 (2) | 8 (2) | 1 (0) | 58 (19) |
| Gram-negative bacteria | 116 (5) | 11 (3) | 21 (4) | 3 (1) | 8 (4) | 8 (2) | 3 (1) | 62 (20) |
| Gram-variable bacteria | 1 (0) | 0 (0) | 0 (0) | 0 (0) | 0 (0) | 0 (0) | 0 (0) | 1 (0) |
| Viruses | 23 (1) | 6 (1) | 3 (1) | 1 (0) | 0 (0) | 7 (1) | 0 (0) | 6 (2) |
| Fungi | 1 (0) | 0 (0) | 0 (0) | 0 (0) | 0 (0) | 0 (0) | 1 (0) | 0 (0) |
| Protozoa | 2 (0) | 0 (0) | 2 (0) | 0 (0) | 0 (0) | 0 (0) | 0 (0) | 0 (0) |

<sup>1</sup>2278 stillbirths had no other condition in the fetus and 23 stillbirths had two other conditions in the fetus.

**Supplementary Table 3.** Main condition in the fetus attributed to death among 2492 stillbirths with complete cause of death determination, by whether placenta specimen was collected, 2016-2023.

| Characteristic | Total<br>N=2492<br>n (%) | Placenta collected<br>N=1224<br>n (%) | Placenta not collected<br>N=1268<br>n (%) |
| --- | --- | --- | --- |
| **Main condition in the fetus** |  |  |  |
| Intrauterine hypoxia | 1864 (75) | 955 (78) | 909 (72) |
| Congenital birth defects | 218 (9) | 79 (6) | 139 (11) |
| Congenital infection | 200 (8) | 106 (9) | 94 (7) |
| Intra-uterine growth restriction | 32 (1) | 17 (1) | 15 (1) |
| Preterm birth complications | 6 (0) | 2 (0) | 4 (0) |
| Aspiration syndrome | 2 (0) | 0 (0) | 2 (0) |
| Birth trauma | 1 (0) | 1 (0) | 0 (0) |
| Cancer | 1 (0) | 0 (0) | 1 (0) |
| Poisoning | 1 (0) | 0 (0) | 1 (0) |
| Other | 17 (1) | 10 (1) | 7 (1) |
| Undetermined | 150 (6) | 54 (4) | 96 (8) |
| **Main maternal condition** |  |  |  |
| Placental complications | 446 (18) | 273 (22) | 173 (14) |
| Maternal hypertension | 414 (17) | 205 (17) | 209 (16) |
| Chorioamnionitis and membrane complications | 173 (7) | 136 (11) | 37 (3) |
| Obstructed labor and fetal malpresentation | 143 (6) | 43 (4) | 100 (8) |
| Other factor (M3) | 133 (5) | 64 (5) | 69 (5) |
| Umbilical cord complications | 105 (4) | 49 (4) | 56 (4) |
| Other factor (M4) | 64 (3) | 42 (3) | 22 (2) |
| Multiple gestation | 59 (2) | 19 (2) | 40 (3) |
| Maternal infection | 58 (2) | 31 (3) | 27 (2) |
| Premature rupture of membranes | 34 (1) | 12 (1) | 22 (2) |
| Maternal diabetes | 31 (1) | 20 (2) | 11 (1) |
| Uterine fluid disorders | 23 (1) | 15 (1) | 8 (1) |
| Prolonged pregnancy | 18 (1) | 9 (1) | 9 (1) |
| Uterine rupture | 18 (1) | 9 (1) | 9 (1) |
| Other factor (Other) | 15 (1) | 6 (0) | 9 (1) |
| Other factor (M2) | 11 (0) | 4 (0) | 7 (1) |
| HIV | 9 (0) | 5 (0) | 4 (0) |
| Maternal injury and accident | 7 (0) | 4 (0) | 3 (0) |
| Preterm labor or delivery | 6 (0) | 2 (0) | 4 (0) |
| Cervical insufficiency and pelvic anomalies | 4 (0) | 1 (0) | 3 (0) |
| Maternal nutritional disorders | 4 (0) | 3 (0) | 1 (0) |
| Maternal circulatory and respiratory diseases | 3 (0) | 2 (0) | 1 (0) |
| Sickle cell disorders | 3 (0) | 2 (0) | 1 (0) |
| Anemias | 2 (0) | 0 (0) | 2 (0) |
| Obesity | 2 (0) | 1 (0) | 1 (0) |
| Other endocrine, metabolic, blood, and immune disorders | 2 (0) | 2 (0) | 0 (0) |
| Poisoning | 2 (0) | 0 (0) | 2 (0) |
| Cesarean delivery | 1 (0) | 1 (0) | 0 (0) |
| Heart Diseases | 1 (0) | 1 (0) | 0 (0) |
| Liver disease | 1 (0) | 0 (0) | 1 (0) |
| Maternal medication or toxic exposure | 1 (0) | 1 (0) | 0 (0) |
| Other respiratory disease | 1 (0) | 1 (0) | 0 (0) |
| No condition identified | 698 (28) | 261 (21) | 437 (34) |
| **DeCoDe level of confidence for main condition in the fetus** |  |  |  |
| Confident | 1889 (81) | 933 (81) | 956 (82) |
| Probable | 222 (10) | 118 (10) | 104 (9) |
| Uncertain | 214 (9) | 102 (9) | 112 (10) |
| **Stillbirth type** |  |  |  |
| Fresh | 1282 (51) | 586 (48) | 696 (55) |
| Macerated | 1120 (45) | 607 (50) | 513 (40) |
| Unknown | 90 (4) | 31 (3) | 59 (5) |
| **Stillbirth timing** |  |  |  |
| Antepartum | 1077 (43) | 602 (49) | 475 (37) |
| Intrapartum | 825 (33) | 353 (29) | 472 (37) |
| Unknown | 590 (24) | 269 (22) | 321 (25) |

**Supplementary Figure 3:**
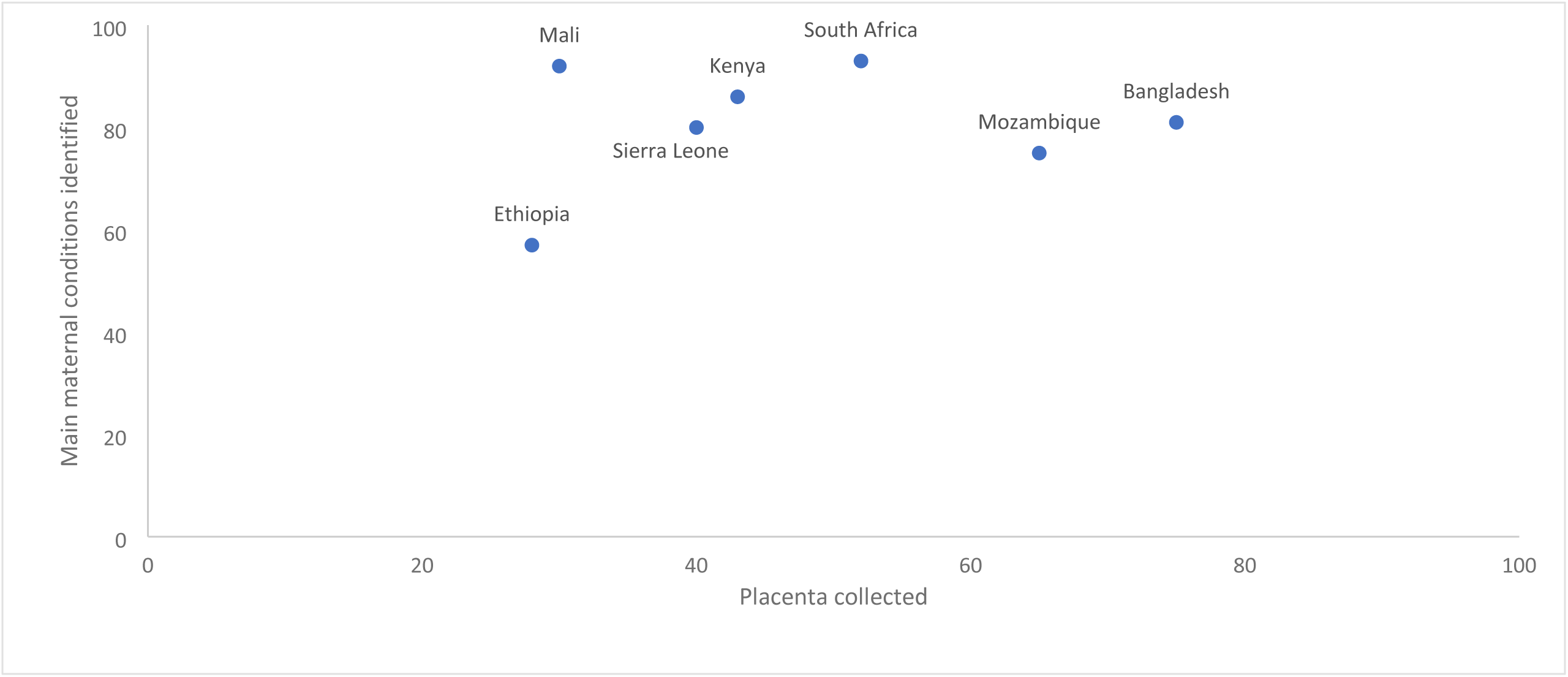
Main maternal conditions identified among 2492 stillbirths and placenta collected by sites, 2016-2023.

**Supplementary Table 4.** Main maternal conditions among stillbirths with intrauterine hypoxia, congenital infection, and congenital birth defects as main condition in the fetus.

| Main maternal condition |  | All Stillbirths<br>N=2492 | Intrauterine hypoxia<br>N=1864<br>n (%) | Congenital birth defects<br>N=218<br>n (%) | Congenital infections<br>N=200<br>n (%) |
| --- | --- | --- | --- | --- | --- |
| ICD-PM Group | Specific |  |  |  |  |
| M1:<br>Complications of<br>placenta, cord<br>and membranes | Placental complications | 446 (18) | 415 (22) | 1 (0) | 7 (4) |
|  | Chorioamnionitis and membrane complications | 173 (7) | 92 (5) | 2 (1) | 73 (36) |
|  | Umbilical cord complications | 105 (4) | 103 (6) | 0 (0) | 1 (0) |
| M2:<br>Complications of<br>pregnancy | Multiple gestation | 59 (2) | 44 (2) | 4 (2) | 0 (0) |
|  | Premature rupture of membranes | 34 (1) | 23 (1) | 0 (0) | 11 (6) |
|  | Uterine fluid disorders | 23 (1) | 20 (1) | 1 (0) | 2 (1) |
|  | Other factor (M2) | 11 (0) | 5 (0) | 0 (0) | 1 (0) |
|  | Cervical insufficiency and pelvic anomalies | 4 (0) | 2 (0) | 0 (0) | 2 (1) |
| M3: Other<br>complications of<br>labor and<br>delivery | Obstructed labor and fetal malpresentation | 143 (6) | 133 (7) | 6 (3) | 2 (1) |
|  | Other factor (M3) | 133 (5) | 129 (7) | 1 (0) | 3 (2) |
|  | Prolonged pregnancy | 18 (1) | 17 (1) | 0 (0) | 0 (0) |
|  | Uterine rupture | 18 (1) | 17 (1) | 1 (0) | 0 (0) |
|  | Preterm labor or delivery | 6 (0) | 6 (0) | 0 (0) | 0 (0) |
|  | Cesarean delivery | 1 (0) | 1 (0) | 0 (0) | 0 (0) |
| M4: Medical and<br>surgical<br>condition | Maternal hypertension | 414 (17) | 382 (20) | 3 (1) | 13 (6) |
|  | Other factor (M4) | 64 (3) | 54 (3) | 0 (0) | 6 (3) |
|  | Maternal infection | 58 (2) | 30 (2) | 1 (0) | 25 (12) |
|  | Maternal diabetes | 31 (1) | 22 (1) | 3 (1) | 4 (2) |
|  | HIV | 9 (0) | 6 (0) | 0 (0) | 3 (2) |
|  | Maternal injury and accident | 7 (0) | 6 (0) | 0 (0) | 0 (0) |
|  | Maternal nutritional disorders | 4 (0) | 3 (0) | 1 (0) | 0 (0) |
|  | Maternal circulatory and respiratory diseases | 3 (0) | 3 (0) | 0 (0) | 0 (0) |
|  | Sickle cell disorders | 3 (0) | 2 (0) | 0 (0) | 0 (0) |
|  | Anemias | 2 (0) | 1 (0) | 0 (0) | 0 (0) |
|  | Obesity | 2 (0) | 2 (0) | 0 (0) | 0 (0) |
|  | Other endocrine, metabolic, blood, and immune disorders | 2 (0) | 2 (0) | 0 (0) | 0 (0) |
|  | Heart Diseases | 1 (0) | 1 (0) | 0 (0) | 0 (0) |
|  | Liver disease | 1 (0) | 1 (0) | 0 (0) | 0 (0) |
|  | Other respiratory disease | 1 (0) | 1 (0) | 0 (0) | 0 (0) |
| Other maternal conditions<sup>1</sup> | Other maternal conditions<sup>1</sup> | 18 (1) | 9 (0) | 3 (1) | 3 (2) |
| No maternal condition identified | No maternal condition identified | 698 (28) | 332 (18) | 191 (88) | 44 (22) |

<sup>1</sup>Includes M35.9, O04, P04.1, P96.4, QA43.Y, QA49, R45.6, T65.9, X68, Y66, Z35.3, Z35.5, Z35.6, Z55.9, Z87.5

**Supplementary Table 5:**
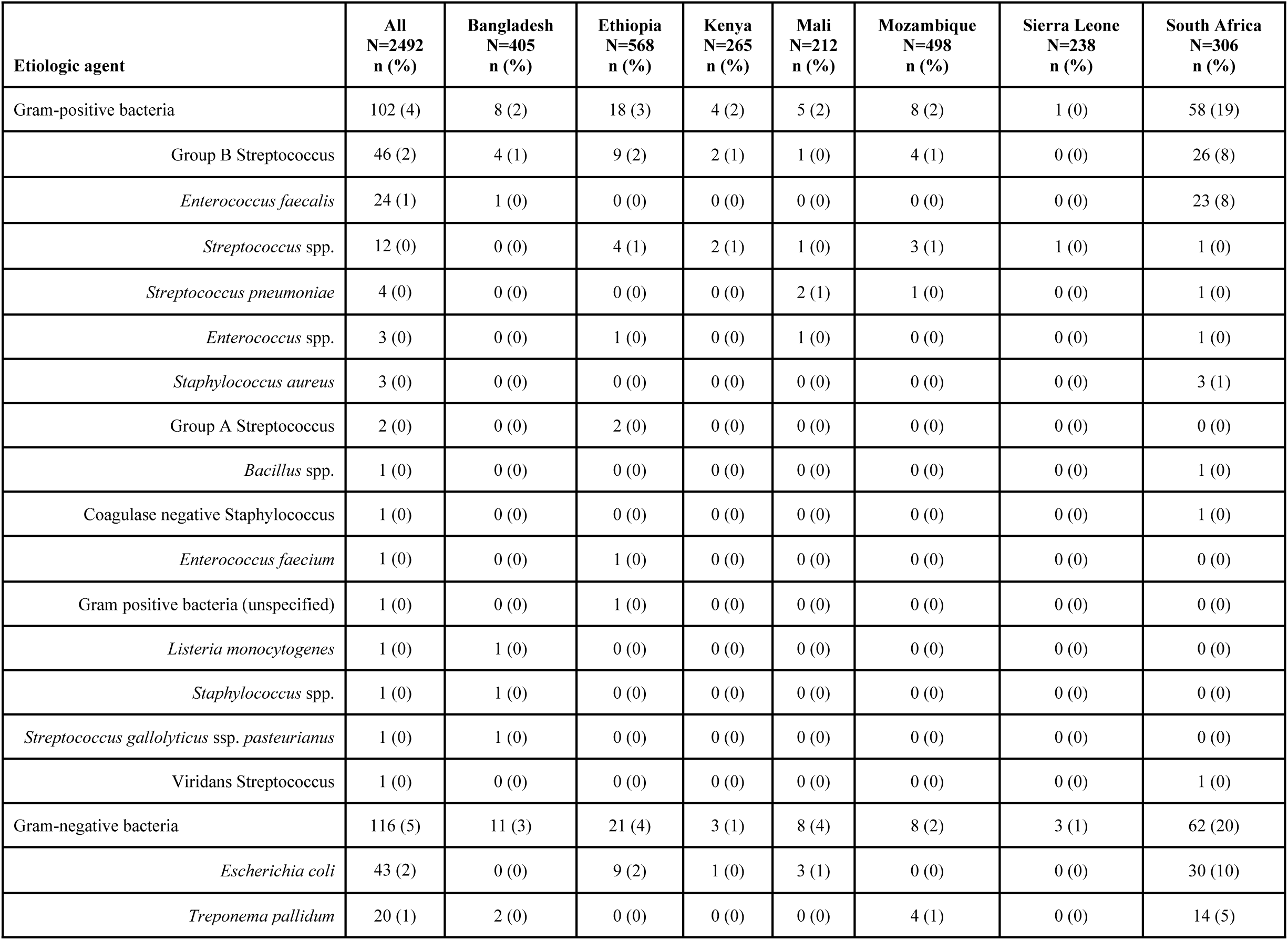
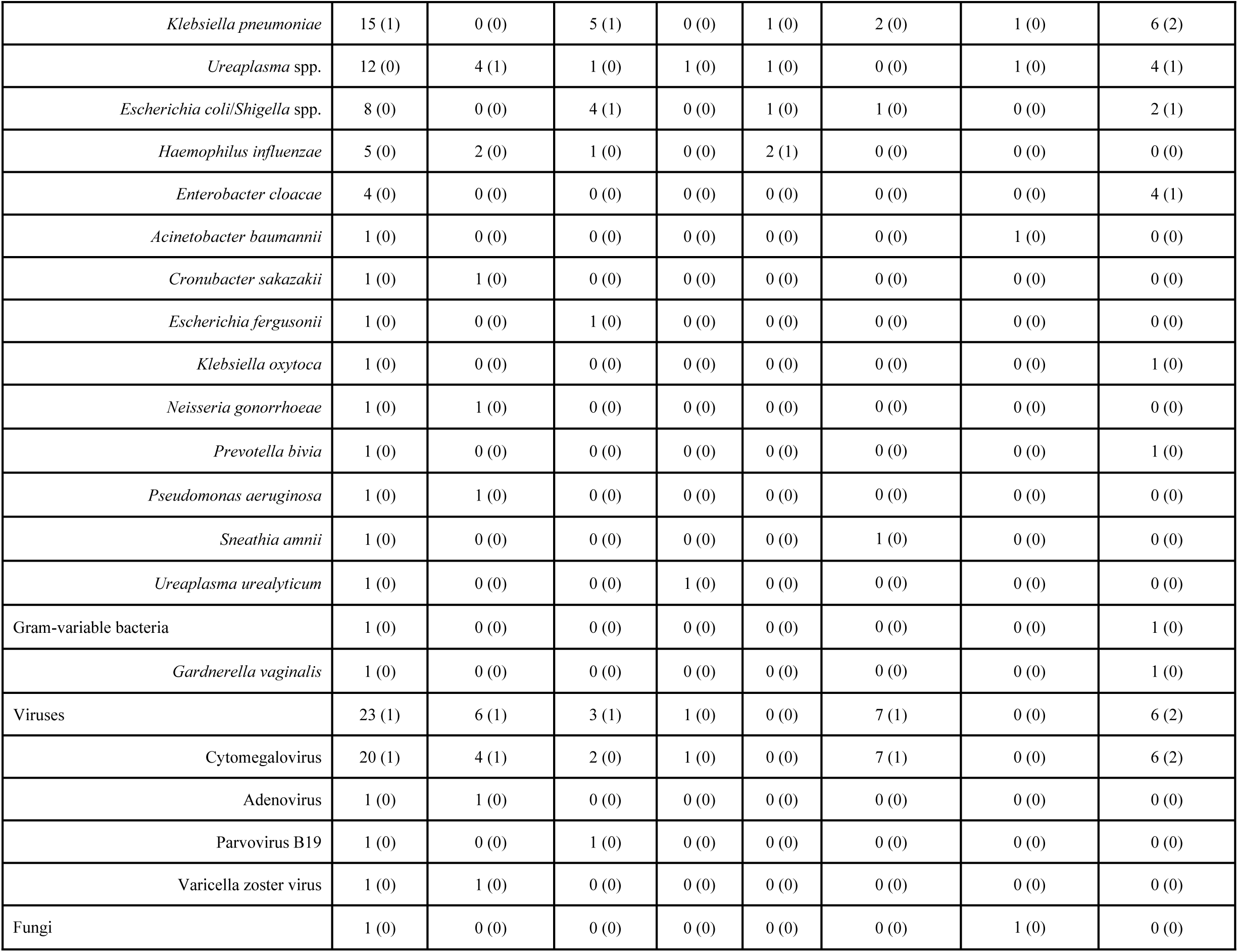
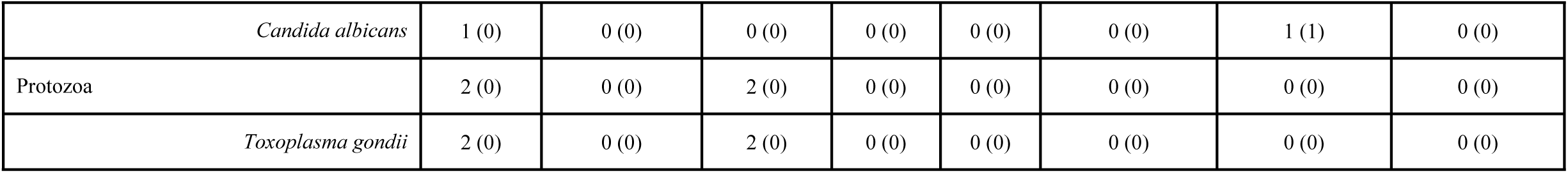
Etiologic agents among stillbirths, all CHAMPS sites, 2016-2023.

**Supplementary Table 6:**
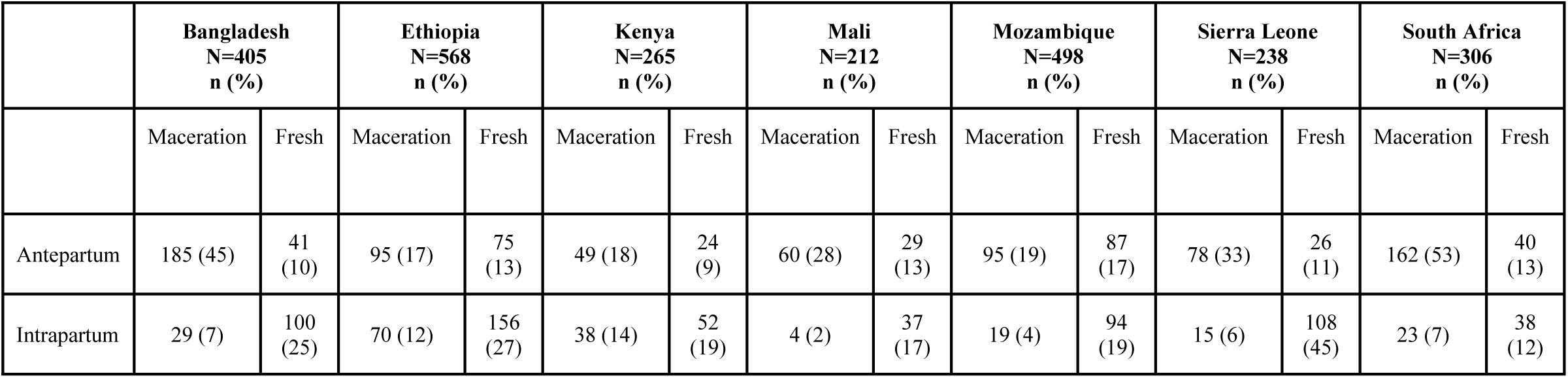
Type and timing of stillbirth by CHAMPS sites, 2016-2023.

|  | **Bangladesh**<br>N=405<br>n (%) |  | **Ethiopia**<br>N=568<br>n (%) |  | **Kenya**<br>N=265<br>n (%) |  | **Mali**<br>N=212<br>n (%) |  | **Mozambique**<br>N=498<br>n (%) |  | **Sierra Leone**<br>N=238<br>n (%) |  | **South Africa**<br>N=306<br>n (%) |  |
| --- | --- | --- | --- | --- | --- | --- | --- | --- | --- | --- | --- | --- | --- | --- |
|  | Maceration | Fresh | Maceration | Fresh | Maceration | Fresh | Maceration | Fresh | Maceration | Fresh | Maceration | Fresh | Maceration | Fresh |
| Antepartum | 185 (45) | 41 (10) | 95 (17) | 75 (13) | 49 (18) | 24 (9) | 60 (28) | 29 (13) | 95 (19) | 87 (17) | 78 (33) | 26 (11) | 162 (53) | 40 (13) |
| Intrapartum | 29 (7) | 100 (25) | 70 (12) | 156 (27) | 38 (14) | 52 (19) | 4 (2) | 37 (17) | 19 (4) | 94 (19) | 15 (6) | 108 (45) | 23 (7) | 38 (12) |

**Supplementary Table 7:** Main condition in the fetus and main maternal condition by type of stillbirth, 2016-2023.

|  | **Total**<br>N=2492<br>n (%) | **Antepartum**<br>N=1077<br>n (%) | **Intrapartum**<br>N=825<br>n (%) | **Unknown**<br>N=590<br>n (%) |
| --- | --- | --- | --- | --- |
| Main condition in the fetus |  |  |  |  |
| Intrauterine hypoxia | 1864 (75) | 801 (74) | 650 (79) | 413 (70) |
| Congenital birth defects | 218 (9) | 69 (6) | 86 (10) | 63 (11) |
| Congenital infection | 200 (8) | 102 (9) | 48 (6) | 50 (8) |
| Intrauterine growth restriction | 32 (1) | 15 (1) | 8 (1) | 9 (2) |
| Preterm birth complications | 6 (0) | 3 (0) | 1 (0) | 2 (0) |
| Aspiration syndrome | 2 (0) | 0 (0) | 0 (0) | 2 (0) |
| Birth trauma | 1 (0) | 0 (0) | 1 (0) | 0 (0) |
| Cancer | 1 (0) | 1 (0) | 0 (0) | 0 (0) |
| Poisoning | 1 (0) | 1 (0) | 0 (0) | 0 (0) |
| Other | 17 (1) | 6 (1) | 7 (1) | 4 (1) |
| Undetermined | 150 (6) | 79 (7) | 24 (3) | 47 (8) |
| Main maternal condition (ICD-PM group) |  |  |  |  |
| M1: Complications of placenta, cord and membranes | 724 (29) | 345 (32) | 216 (26) | 163 (28) |
| M2: Complications of pregnancy | 131 (5) | 59 (5) | 44 (5) | 28 (5) |
| M3: Other complications of labor and delivery | 319 (13) | 43 (4) | 213 (26) | 63 (11) |
| M4: Medical and surgical condition | 602 (24) | 331 (31) | 156 (19) | 115 (19) |
| Other maternal condition | 18 (1) | 13 (1) | 4 (0) | 1 (0) |
| No maternal condition identified | 698 (28) | 286 (27) | 192 (23) | 220 (37) |
| Fetal heartbeat during labor |  |  |  |  |
| Present | 342 (14) | 57 (5) | 202 (24) | 83 (14) |
| Absent | 783 (31) | 398 (37) | 249 (30) | 136 (23) |
| Data missing | 1367 (55) | 622 (58) | 374 (45) | 371 (63) |
| Stillbirth type |  |  |  |  |
| Fresh | 1282 (51) | 322 (30) | 585 (71) | 375 (64) |
| Macerated | 1120 (45) | 724 (67) | 198 (24) | 198 (33) |
| Unknown | 90 (4) | 31 (3) | 42 (5) | 17 (3) |

**Supplementary Table 8:** Availability of maternal medical record among 2492 stillbirths, 2016-2023.

|  | All<br>N=2492<br>n (%) | Bangladesh<br>N=405<br>n (%) | Ethiopia<br>N=568<br>n (%) | Kenya<br>N=265<br>n (%) | Mali<br>N=212<br>n (%) | Mozambique<br>N=498<br>n (%) | Sierra<br>Leone<br>N=238<br>n (%) | South<br>Africa<br>N=306<br>n (%) |
| --- | --- | --- | --- | --- | --- | --- | --- | --- |
| **Maternal medical record** |  |  |  |  |  |  |  |  |
| Has maternal condition | 1794 (72) | 327 (81) | 266 (47) | 216 (82) | 180 (85) | 357 (72) | 182 (76) | 266 (87) |
| Has medical record and no maternal condition | 576 (23) | 78 (19) | 223 (39) | 49 (18) | 24 (11) | 112 (22) | 55 (23) | 35 (11) |
| No medical record and no maternal condition | 122 (5) | 0 (0) | 79 (14) | 0 (0) | 8 (4) | 29 (6) | 1 (0) | 5 (2) |

**Supplementary Table 9.** Main maternal conditions among stillbirths by small-for-gestational age.

| Main maternal condition |  | All Stillbirths<br>N=2492 | Small-for-gestational age<br>N=625<br>n (%) | Not small-for-gestational age<br>N=1101<br>n (%) | Unknown<br>N=766<br>n (%) |
| --- | --- | --- | --- | --- | --- |
| ICD-PM Group | Specific |  |  |  |  |
| M1: Complications of placenta, cord and membranes | Placental complications | 446 (18) | 129 (21) | 209 (19) | 108 (14) |
|  | Chorioamnionitis and membrane complications | 173 (7) | 45 (7) | 89 (8) | 39 (5) |
|  | Umbilical cord complications | 105 (4) | 19 (3) | 53 (5) | 33 (4) |
| M2: Complications of pregnancy | Multiple gestation | 59 (2) | 18 (3) | 14 (1) | 27 (4) |
|  | Premature rupture of membranes | 34 (1) | 3 (0) | 18 (2) | 13 (2) |
|  | Uterine fluid disorders | 23 (1) | 12 (2) | 7 (1) | 4 (1) |
|  | Other factor (M2) | 11 (0) | 0 (0) | 7 (1) | 4 (1) |
|  | Cervical insufficiency and pelvic anomalies | 4 (0) | 1 (0) | 2 (0) | 1 (0) |
| M3: Other complications of labor and delivery | Obstructed labor and fetal malpresentation | 143 (6) | 20 (3) | 75 (7) | 48 (6) |
|  | Other factor (M3) | 133 (5) | 18 (3) | 76 (7) | 39 (5) |
|  | Prolonged pregnancy | 18 (1) | 7 (1) | 5 (0) | 6 (1) |
|  | Uterine rupture | 18 (1) | 0 (0) | 14 (1) | 4 (1) |
|  | Preterm labor or delivery | 6 (0) | 1 (0) | 3 (0) | 2 (0) |
|  | Cesarean delivery | 1 (0) | 1 (0) | 0 (0) | 0 (0) |
| M4: Medical and surgical condition | Maternal hypertension | 414 (17) | 141 (23) | 185 (17) | 88 (11) |
|  | Other factor (M4) | 64 (3) | 15 (2) | 40 (4) | 9 (1) |
|  | Maternal infection | 58 (2) | 18 (3) | 31 (3) | 9 (1) |
|  | Maternal diabetes | 31 (1) | 6 (1) | 17 (2) | 8 (1) |
|  | HIV | 9 (0) | 1 (0) | 7 (1) | 1 (0) |
|  | Maternal injury and accident | 7 (0) | 3 (0) | 3 (0) | 1 (0) |
|  | Maternal nutritional disorders | 4 (0) | 3 (0) | 0 (0) | 1 (0) |
|  | Maternal circulatory and respiratory diseases | 3 (0) | 2 (0) | 1 (0) | 0 (0) |
|  | Sickle cell disorders | 3 (0) | 1 (0) | 2 (0) | 0 (0) |
|  | Anemias | 2 (0) | 0 (0) | 2 (0) | 0 (0) |
|  | Obesity | 2 (0) | 0 (0) | 2 (0) | 0 (0) |
|  | Other endocrine, metabolic, blood, and immune disorders | 2 (0) | 1 (0) | 1 (0) | 0 (0) |
|  | Heart Diseases | 1 (0) | 0 (0) | 0 (0) | 1 (0) |
|  | Liver disease | 1 (0) | 0 (0) | 0 (0) | 1 (0) |
|  | Other respiratory disease | 1 (0) | 0 (0) | 0 (0) | 1 (0) |
| Other maternal conditions<sup>1</sup> | Other maternal conditions<sup>1</sup> | 18 (1) | 5 (1) | 8 (1) | 5 (1) |
| No maternal condition identified | No maternal condition identified | 698 (28) | 155 (25) | 230 (21) | 313 (41) |

<sup>1</sup>Includes M35.9, O04, P04.1, P96.4, QA43.Y, QA49, R45.6, T65.9, X68, Y66, Z35.3, Z35.5, Z35.6, Z55.9, Z87.5

**Supplementary Table 10.** Recommendations for 1808 preventable stillbirth cases, by CHAMPS site, 2016-2023.

| Recommendations<sup>1</sup> | Total<br>N=1808<br>n (%) | Bangladesh<br>N=364<br>n (%) | Ethiopia<br>N=470<br>n (%) | Kenya<br>N=250<br>n (%) | Mali<br>N=146<br>n (%) | Mozambique<br>N=280<br>n (%) | Sierra Leone<br>N=206<br>n (%) | South Africa<br>N=92<br>n (%) |
| --- | --- | --- | --- | --- | --- | --- | --- | --- |
| Improved family planning | 305 (17) | 24 (7) | 247 (53) | 18 (7) | 2 (1) | 5 (2) | 7 (3) | 2 (2) |
| Improved antenatal care and obstetric care and management | 1532 (85) | 303 (83) | 392 (83) | 234 (94) | 108 (74) | 272 (97) | 149 (72) | 74 (80) |
| Improved infection prevention and control | 74 (4) | 5 (1) | 29 (6) | 15 (6) | 1 (1) | 21 (8) | 2 (1) | 1 (1) |
| Improved HIV prevention and control | 141 (8) | 6 (2) | 46 (10) | 60 (24) | 0 (0) | 3 (1) | 24 (12) | 2 (2) |
| Improved health-seeking behavior | 491 (27) | 56 (15) | 267 (57) | 93 (37) | 9 (6) | 13 (5) | 36 (17) | 17 (18) |
| Improved nutritional support | 100 (6) | 5 (1) | 92 (20) | 1 (0) | 0 (0) | 1 (0) | 1 (0) | 0 (0) |
| Improved vaccinations | 6 (0) | 1 (0) | 2 (0) | 0 (0) | 1 (1) | 2 (1) | 0 (0) | 0 (0) |
| Improved transport system | 66 (4) | 0 (0) | 42 (9) | 4 (2) | 4 (3) | 13 (5) | 2 (1) | 1 (1) |
| Other recommendation | 351<br>(19) | 66 (18) | 196<br>(42) | 55<br>(22) | 4 (3) | 5 (2) | 22 (11) | 3 (3) |

<sup>1</sup>Single case can have multiple recommendations. Includes deaths that are preventable given current situation or preventable given ideal situation.

## Notes

### Author Declarations

. Ethics committees in each CHAMPS site and Emory University approved of the study protocols. (https://champshealth.org/resources/protocols/)

## References

1. United Nations Inter-agency Group for Child Mortality Estimation. Never Forgotten: The situation of stillbirths around the globe, 2023.

2. Blencowe H, Cousens S, Jassir FB, et al. National, regional, and worldwide estimates of stillbirth rates in 2015, with trends from 2000: a systematic analysis. Lancet Glob Health 2016; 4(2): e98–e108.

3. United Nations Inter-agency Group for Child Mortality Estimation. Levels and Trends in Child Mortality: Report 2022, 2023.

4. Lawn JE, Blencowe H, Waiswa P, et al. Stillbirths: rates, risk factors, and acceleration towards 2030. Lancet 2016; 387(10018): 587–603.

5. Madhi SA, Pathirana J, Baillie V, et al. An Observational Pilot Study Evaluating the Utility of Minimally Invasive Tissue Sampling to Determine the Cause of Stillbirths in South African Women. Clin Infect Dis 2019; 69(Suppl 4): S342–S50.

6. Reinebrant HE, Leisher SH, Coory M, et al. Making stillbirths visible: a systematic review of globally reported causes of stillbirth. BJOG 2018; 125(2): 212–24.

7. Mensah Abrampah NA, Okwaraji YB, You D, et al. Global Stillbirth Policy Review – Outcomes And Implications Ahead of the 2030 Sustainable Development Goal Agenda. International Journal of Health Policy and Management 2023; 12(Issue 1): 1–11.

8. Global, regional, and national stillbirths at 20 weeks’ gestation or longer in 204 countries and territories, 1990-2021: findings from the Global Burden of Disease Study 2021. Lancet 2024; 404(10466): 1955–88.

9. Raghunathan PL, Madhi SA, Breiman RF. Illuminating Child Mortality: Discovering Why Children Die. Clinical Infectious Diseases 2019; 69(Supplement_4): S257–S9.

10. Taylor AW, Blau DM, Bassat Q, et al. Initial findings from a novel population-based child mortality surveillance approach: a descriptive study. The Lancet Global Health 2020; 8(7): e909–e19.

11. Salzberg NT, Sivalogan K, Bassat Q, et al. Mortality Surveillance Methods to Identify and Characterize Deaths in Child Health and Mortality Prevention Surveillance Network Sites. Clin Infect Dis 2019; 69(Suppl 4): S262–S73.

12. Blevins J, O’Mara Sage E, Kone A, et al. Using Participatory Workshops to Assess Alignment or Tension in the Community for Minimally Invasive Tissue Sampling Prior to Start of Child Mortality Surveillance: Lessons From 5 Sites Across the CHAMPS Network. Clinical Infectious Diseases 2019; 69(Supplement_4): S280–S90.

13. O’Mara Sage E, Munguambe KR, Blevins J, et al. Investigating the Feasibility of Child Mortality Surveillance With Postmortem Tissue Sampling: Generating Constructs and Variables to Strengthen Validity and Reliability in Qualitative Research. Clin Infect Dis 2019; 69(Suppl 4): S291–S301.

14. Rakislova N, Fernandes F, Lovane L, et al. Standardization of Minimally Invasive Tissue Sampling Specimen Collection and Pathology Training for the Child Health and Mortality Prevention Surveillance Network. Clin Infect Dis 2019; 69(Suppl 4): S302–S10.

15. Martines RB, Ritter JM, Gary J, et al. Pathology and Telepathology Methods in the Child Health and Mortality Prevention Surveillance Network. Clin Infect Dis 2019; 69(Suppl 4): S322–S32.

16. Diaz MH, Waller JL, Theodore MJ, et al. Development and Implementation of Multiplex TaqMan Array Cards for Specimen Testing at Child Health and Mortality Prevention Surveillance Site Laboratories. Clin Infect Dis 2019; 69(Suppl 4): S311–S21.

17. Villar J, Cheikh Ismail L, Victora CG, et al. International standards for newborn weight, length, and head circumference by gestational age and sex: the Newborn Cross-Sectional Study of the INTERGROWTH-21st Project. Lancet 2014; 384(9946): 857–68.

18. Blau DM, Caneer JP, Philipsborn RP, et al. Overview and Development of the Child Health and Mortality Prevention Surveillance Determination of Cause of Death (DeCoDe) Process and DeCoDe Diagnosis Standards. Clin Infect Dis 2019; 69(Suppl 4): S333–S41.

19. Madrid L, Vyas KJ, Kancherla V, et al. Neural tube defects as a cause of death among stillbirths, infants, and children younger than 5 years in sub-Saharan Africa and southeast Asia: an analysis of the CHAMPS network. The Lancet Global Health 2023; 11(7): e1041–e52.

20. Schutte AE, Srinivasapura Venkateshmurthy N, Mohan S, Prabhakaran D. Hypertension in Low- and Middle-Income Countries. Circulation Research 2021; 128(7): 808–26.

21. Wang W, Xie X, Yuan T, et al. Epidemiological trends of maternal hypertensive disorders of pregnancy at the global, regional, and national levels: a population-based study. BMC Pregnancy and Childbirth 2021; 21(1).

22. Tannor EK, Nyarko OO, Adu-Boakye Y, et al. Prevalence of Hypertension in Ghana: Analysis of an Awareness and Screening Campaign in 2019. Clin Med Insights Cardiol 2022; 16: 11795468221120092.

23. Berhe AK, Ilesanmi AO, Aimakhu CO, Bezabih AM. Awareness of pregnancy induced hypertension among pregnant women in Tigray Regional State, Ethiopia. Pan Afr Med J 2020; 35: 71.

24. Finlayson K, Downe S. Why Do Women Not Use Antenatal Services in Low- and Middle-Income Countries? A Meta-Synthesis of Qualitative Studies. PLoS Medicine 2013; 10(1): e1001373.

25. Billah SM, Khan ANS, Rokonuzzaman SM, et al. Competency of health workers in detecting and managing gestational hypertension, pre-eclampsia, severe pre-eclampsia and eclampsia during antenatal check-ups in primary care health facilities in Bangladesh: a cross-sectional study. BMJ Open 2021; 11(7): e046638.

26. Rawlins B, Plotkin M, Rakotovao JP, et al. Screening and management of pre-eclampsia and eclampsia in antenatal and labor and delivery services: findings from cross-sectional observation studies in six sub-Saharan African countries. BMC Pregnancy and Childbirth 2018; 18(1).

27. Oyelese Y, Ananth CV. Placental abruption. Obstet Gynecol 2006; 108(4): 1005–16.

28. Cutland CL, Schrag SJ, Zell ER, et al. Maternal HIV infection and vertical transmission of pathogenic bacteria. Pediatrics 2012; 130(3): e581–90.

29. Madhi SA, Briner C, Maswime S, et al. Causes of stillbirths among women from South Africa: a prospective, observational study. The Lancet Global Health 2019; 7(4): e503–e12.

30. MRC Vitamin Study Research Group. Prevention of neural tube defects: results of the Medical Research Council Vitamin Study. Lanc*e*t 1991; 338(8760): 131–7.

31. Lawn JE, Ohuma EO, Bradley E, et al. Small babies, big risks: global estimates of prevalence and mortality for vulnerable newborns to accelerate change and improve counting. Lancet 2023; 401(10389): 1707–19.

32. Reddy UM, Page GP, Saade GR, et al. Karyotype versus microarray testing for genetic abnormalities after stillbirth. N Engl J Med 2012; 367(23): 2185–93.

